# Low-Cost, High-Volume Health Services Contribute the Most to Unnecessary Health Spending due to Low-Value Care in Japan

**DOI:** 10.1101/2025.08.21.25334207

**Authors:** Atsushi Miyawaki, John N. Mafi, Tsuguya Fukui, Yuya Kimura, Daiki Kobayashi, Sara Odawara, Kazuhiro Abe, Rei Goto, Yusuke Tsugawa

**Author notes:** **Corresponding Author:** Atsushi Miyawaki, MD, PhD, Public Health Research Group, Institute of Medicine, University of Tsukuba Address: 1-1-1 Tennodai, Tsukuba, Ibaraki 305-8574, Japan.

## Abstract

**Importance:** As healthcare costs continue to rise, high-income countries—including Japan—face the urgent task of reducing healthcare spending incurred by low-value care. However, evidence is limited as to which low-value care services contribute most to unnecessary healthcare spending outside of the United States.

**Objective:** To identify which low-value care services contribute the most to unnecessary healthcare spending in Japan.

**Design, Setting, and Participants:** The cross-sectional study of all beneficiaries using a population-based claims database from April 1, 2022, to March 31, 2023, encompassing all age groups, reflecting approximately 2% of the total Japanese population.

**Main Outcomes and Measures:** We identified 52 low-value care services based on clinical evidence, and examined their contributions to healthcare spending using two versions of claims-based measures with different sensitivities and specificities (broader and narrower definitions). Each service was categorized into four groups based on its average per-service price: very low (<1,000 Japanese yen [JPY] = 8 US dollars [USD] in 2022), low (1,000–9,999 JPY), medium (10,000–99,999 JPY), or high (≥100,000 JPY).

**Results:** Among 1,923,484 beneficiaries (mean [SD] age 58.6 [23.5] years; 52.7% female), we identified 3.1 million (narrower definition) to 3.7 million (broader definition) episodes of low-value care services (1.6–1.9 per capita), with 36–40% of patients receiving at least one low-value care service. These services accounted for 0.7–1.0% of total healthcare spending, amounting to 207–331 billion JPY (1.7–2.6 billion USD) when extrapolated nationwide with adjustments for age, sex, and region. When applying narrower definitions, over 99% of low-value care episodes involved very-low-cost or low-cost services, which accounted for 67% of unnecessary healthcare spending—far exceeding the 33% attributed to medium-cost or high-cost services.

**Conclusion and Relevance:** Over one in three Japanese individuals received low-value care during 2022-2023, contributing to 0.7–1.0% of total healthcare spending. Among these services, low-cost services contributed to virtually all low-value care utilization and over two-thirds of unnecessary healthcare spending. Compared to focusing solely on high-cost services, targeting the reduction of frequently performed, lower-cost services may be a more effective strategy for reducing wasteful spending.

**KEY POINTS:** 

**Question:** Which low-value care services—low-cost or high-cost—contribute most to unnecessary healthcare spending in Japan?

**Findings:** In a cross-sectional study of nearly two million beneficiaries examining 52 low-value care services, over one-third received at least one such service during a one-year period, accounting for 0.7–1.0% of total healthcare spending. More than 99% of episodes were very-low- or low-cost services, accounting for over two-thirds of low-value care spending, exceeding spending from medium- and high-cost services.

**Meaning:** Focusing on frequently performed, lower-cost services may better reduce wasteful healthcare spending than targeting only high-cost services.

## INTRODUCTION

As healthcare spending growth continues to outpace inflation, many high-income countries are facing the urgent task of curbing unsustainable spending growth. Reducing low-value care (LVC)—healthcare services that provide little or no net clinical benefit in specific scenarios^1–3^—is appealing to policymakers because it can eliminate wasteful spending without compromising the quality of care and patients’ health outcomes. Reducing spending on LVC services has the potential to directly curb unnecessary healthcare spending, improve care quality and patient safety by minimizing overdiagnosis and overtreatment, and lead to better population health outcomes by reallocating healthcare resources toward high-value services.^4^

Despite the publication of numerous guidelines, a global Choosing Wisely campaign, and decades of attention to this issue, spending on LVC services remains persistent.^5–8^ Several studies have reported problems due to the use of high-cost LVC services^9,10^—such as knee arthroscopy among patients with osteoarthritis—and their associated spending.^11–13^ However, existing research has also shown that even low-cost LVC can amount to considerable unnecessary healthcare spending when provided at scale.^14,15^ High- and low-cost services may differ in several respects, including the setting in which care is delivered (hospitals vs. clinics), the specialty of the physician providing the service (specialists vs. generalists), and the characteristics of the patient populations they target (e.g., individuals with severe vs. minor illnesses). Identifying which LVC services contribute more significantly to unnecessary healthcare spending is essential for prioritizing effective strategies to reduce their use.

However, evidence is limited regarding the relative contributions of high-cost and low-cost services to unnecessary healthcare spending. While existing research on this topic suggests that spending on low-cost LVC services exceeds that of high-cost LVC services,^16–18^ these studies are confined to the United States (US), and it is unclear whether this finding can be generalized to countries with different healthcare systems, access to physician services, and payment models.

LVC represents a critical public health issue in Japan, as it does in the US Japan faces the dual challenge of maintaining fiscal sustainability while ensuring patient safety in the context of a rapidly aging population. The widespread coverage of numerous LVC services under social insurance plans,^7,19^ combined with limited perceptions of LVC provision among physicians,^20^ can lead to higher healthcare spending and an increased risk of patient exposure to its adverse effects. In this context, using a nationwide health insurance claims database in Japan, we examined how high-cost and low-cost LVC services contribute to unnecessary healthcare spending.

## METHODS

### Health Systems in Japan

Japan’s health system is characterized by predominantly private clinics and hospitals financed by a combination of social health insurance and cost-sharing from patients. Japanese citizens are required by law to enroll in one of the social health insurance plans (i.e., either employment-based or residence-based insurance), and regardless of the plan, beneficiaries are covered under standardized benefits, such as the uniform fee schedules, the same co-insurance rates (10–30%, varying by age categories), out-of-pocket maximum (covered by the catastrophic health insurance program), and freedom to choose any hospital or clinic (similar to Preferred Provider Organization plans in the US, although some tertiary hospitals charge an additional “first visit fee” for patients without a referral letter from a primary care physician). Insurance benefits are also standardized and include all healthcare services except for preventive services (which are financed using general tax revenues) and long-term care (covered under long-term care insurance). The majority of outpatient services are reimbursed through the fee-for-service model. The majority of inpatient care at large acute care hospitals is paid through a bundled payment called the Diagnosis Procedure Combination, which is a per diem payment system based on diagnosis and procedures (a modified version of the diagnosis-related group in the US). While these features ensure broad access to care, they may also create incentives for clinics or hospitals to increase the volume of physician visits and diagnostic tests.^21^ In turn, this may facilitate the widespread provision of LVC especially in outpatient settings and influence the cost distribution of services that may explain unnecessary spending.

### Data Sources

We used data obtained from the DeSC claims database, a nationwide health insurance claims database compiled and maintained by DeSC Healthcare, Inc. (Tokyo, Japan).^22^ ^23,24^ The dataset, extracted in September 2024, included approximately 2.3 million insured individuals as of April 2023, representing about 2% of the total Japanese population. The database includes all age groups and comprises individuals from diverse socioeconomic backgrounds. It includes beneficiaries in several major social health insurance schemes in Japan: the National Health Insurance program (covering unemployed, self-employed, and retired individuals and their dependents aged <75 years); corporate health insurance societies (covering employed individuals and their dependents aged <75 years); and the Late-Stage Medical Care System (covering all individuals aged ≥75 years). Individuals from households receiving public assistance, who constitute approximately 1.6% of the total Japanese population, were not included in this database because their healthcare costs were covered by public funds rather than health insurance. Although the age distribution in the database is slightly older than that of the total Japanese population, the prevalence of major comorbidities is comparable to estimates from a national survey.^25^

The DeSC database includes the beneficiary registry, medical claims, and dispensing claims. The beneficiary registry contains an encrypted personal identifier and beneficiaries’ demographics, insurance type, and region, enabling longitudinal tracking of individuals across multiple health care settings. Medical claims provide detailed information on both outpatient and inpatient care (service dates, diagnosis codes, and corresponding start and end dates). Dispensing claims document prescriptions, specifying drug names, quantities, and prescription/dispensing dates. Diagnosis codes are recorded using the *International Classification of Diseases, 10th Revision* (ICD-10); drug agents are categorized according to the World Health Organization’s *Anatomical Therapeutic Chemical* classification system; and tests and procedures are recorded using the *Japanese Medical Practice Codes*, a collection of standardized codes representing medical procedures, supplies, products, and services used in Japan.

### Study Population

To examine LVC provided in fiscal year 2022 (April 1, 2022, to March 31, 2023), we included all beneficiaries who were continuously enrolled in the DeSC database from April 2022 through March 2023. We required continuous enrollment over a lookback period of one year preceding the fiscal year 2022 and over subsequent cascade periods of one month, as some LVC services required information on medical history from the preceding year and preoperative status within one month.

### LVC Measurement

This study identified 52 LVC services provided in Japan through a review of published clinical literature (**eTable 1** and **eTable 2** in **Supplement File**). We began with 31 LVC services identified and detected in a previous study conducted in acute care hospital settings in Japan.^7^ Next, to conduct a more comprehensive assessment, we updated this list by assembling a panel of physicians from 19 specialties. Each specialist was asked to list services within their specialty that are definitively low-value—defined as having evidence of no clinical benefit, supported by meta-analyses or multiple studies (including randomized controlled trials) showing no efficacy with minimal variability in outcomes—together with the relevant clinical evidence. Two independent physicians on the research team (A.M. and Y.K.), who are experienced in claims-based analysis, reviewed the provided clinical evidence and selected services consistently categorized as “definitely low-value” by both the specialists and the independent reviewers. They then assessed whether each service was measurable using Japanese claims data, excluding those with insufficient information or not covered by the public health insurance system (**eTable 3** in **Supplement File**). This process resulted in the identification of 24 measurable LVC services. Finally, we combined these 24 newly validated services with the original 31 services, and after removing three duplicates, we established a final set of 52 LVC services. Details of these processes are provided in **eMethod 1** in **Supplement File**. Each service and its operational definition are presented in **eTable 1** in **Supplement File**, with corresponding codes provided in **eTable 2** in **Supplement File**.

To address the inherent uncertainty in quantifying LVC services using administrative claims data, we specified two versions of each LVC measure following previous studies:^1,7^ a broader (more sensitive) definition and a narrower (more specific) definition. First, we created the broader definition to include all LVC, which runs the risk of misclassifying appropriate care as low-value. By adding some criteria to this broader definition, we next created the narrower definition to minimize the misclassification of appropriate care; conversely, this runs the risk of missing some LVC. For example, the prescription of antibiotics for patients diagnosed with an acute respiratory infection was classified as low-value under the broader definition. Under the narrower definition, antibiotics were considered low-value only if there was no accompanying diagnosis for which antibiotics may be appropriate. We adopted operational definitions from previous research in Japan,^7^ using a consensus-based approach, with minor modifications for compatibility with the DeSC claims database. For the newly added services, definitions were established through a consensus method involving three physicians experienced in analyzing healthcare administrative data (A.M., Y.K., and Y.T.).

### Spending Calculations

To quantify spending associated with each LVC service, we calculated the total amount paid to healthcare providers (including patients’ out-of-pocket costs). In doing so, we used government-set prices for each service—which are standardized by law in Japan—across geography, clinical settings, and insurers. For 42 of 52 measures, the spending was defined as costs of the detected LVC service itself to avoid overestimation of the spending on LVC. It should be noted that although inpatient services at large hospitals in Japan are reimbursed under a per diem payment system, we followed a previous study^7^ and calculated spending of detected services on a fee-for-service basis, per the Guideline for Healthcare Spending-Effectiveness Evaluation issued by the Japan Central Social Insurance Council.^26^ For the remaining 10 measures, which were all procedural/surgical services, it was not possible to comprehensively specify the many codes that could be relevant to the service. Therefore, for three services often performed in the outpatient setting (e.g., spinal injection for low back pain), all costs incurred during the same day of service were included in the healthcare spending estimates to capture all related costs within the same episode of care (e.g., in the case of spinal injections, we included the costs of the physician’s effort [examinations and procedures] as well as infusion of the drug^6^). For seven services occurring nearly exclusively in the inpatient setting (e.g., carotid endarterectomy), the total cost of the hospitalization was considered as the healthcare spending estimates to capture all related costs within the same episode of care, given that the hospitalization occurred because of the LVC service. Details are provided in **eTable 2** in **Supplement File**.

### Statistical Analysis

First, we described the characteristics of the beneficiaries. Second, we examined the total volume of identified LVC services and the proportion of beneficiaries who received at least one LVC service from April 2022 to March 2023. As was the case with prior research,^16,27^ we primarily presented the narrower set of LVC measures to minimize the risk of misclassifying high-value care services as LVC services, even at the cost of potentially underestimating the prevalence of LVC. We also reported the associated unnecessary healthcare spending. Furthermore, we analyzed the total LVC volume and associated spending by age group (<18 years, 18–64 years, 65–74 years, and ≥75 years). We also presented estimates of national volume and spending on LVC by extrapolating the age-, sex-, and region-specific per capita volume and spending to the national population (see **eMethod 2** in **Supplement File** for details).

Third, we examined how different service price categories (e.g., low-cost vs. high-cost services) contributed to the total volume and spending of LVC. Service price categories were determined based on their average per-service spending as very low (<1,000 Japanese yen [JPY] or 8 US dollars [USD]; 125 JPY=1 USD in 2022), low (1,000–9,999 JPY [8–80 USD]), medium (10,000–99,999 JPY [80–800 USD]), or high (≥100,000 JPY [800 USD]). Average per-service spending was calculated based on the calculated volume and associated spending for each LVC service in the total sample. Furthermore, we identified the top ten LVC services contributing to unnecessary spending in the sample and reported average per-service spending, LVC volume and associated spending, and percentage in grand total LVC spending.

### Sensitivity Analyses

We conducted a few sensitivity analyses. First, to test whether our findings were sensitive to the definition of LVC, we quantified the LVC volume and spending by using the broader set of more sensitive and less specific LVC measures instead of the narrower one and examined the price category distribution of LVC services. Second, we repeated the analyses using alternative price category cutoffs by classifying LVC services into quartiles based on service price, rather than using the cutoffs of 1,000, 10,000, and 100,000 JPY. Finally, as the analytic sample was slightly older than the national population, we reanalyzed the price distribution of LVC based on age-, sex-, and region-adjusted national extrapolations.

### Stratified Analyses

To examine how the price category distribution of LVC services varied by beneficiaries’ age, we examined the proportion of total LVC volume and spending in each of the price categories for different age categories (<18 years, 18–64 years, 65–74 years, and ≥75 years).

The Ethics Committee of the University of Tokyo approved this study and waived written informed consent because we retrospectively analyzed deidentified data. This manuscript follows the STROBE reporting guidelines for observational studies.

## RESULTS

### Volume of and Spending on LVC Services

The analytic sample included 1,923,484 beneficiaries (mean [SD] age, 58.6 [23.5] years; 52.7% female) (**Table 1**). For these beneficiaries, LVC services defined by 52 LVC measures were provided a total of 3,123,618 times from April 2022 to March 2023, corresponding to 1,623.9 times per 1,000 beneficiaries (**Table 2**). We found that 36.2% of beneficiaries (696,190 individuals) received at least one LVC services annually. In this cohort, unnecessary spending on LVC services totaled 5.3 billion JPY (42.6 million USD), corresponding to 2.78 million JPY (22.2 thousand USD) per 1,000 beneficiaries. This accounted for 0.65% of the overall healthcare spending of 823.7 billion JPY (6.6 billion USD) in this cohort. Age-stratified analyses revealed that the LVC spending per 1,000 beneficiaries increased with beneficiary age, from 0.49 million JPY (3,920 USD) among those aged <18 years to 5.50 million JPY (44.0 thousand USD) among those aged ≥75 years. After extrapolating to the national population with age-, sex-, and region-adjustments, we calculated that the annual national volume and spending on LVC in Japan were 138.8 million services and 207.2 billion JPY (1.7 billion USD), respectively.

**Table 1.** Patient characteristics.

|  | **Beneficiaries (N=1,923,484)** |
| --- | --- |
| **Female sex** | 1,014,208 (52.7) |
| **Age** |  |
| <18 years | 162,563 (8.5) |
| 18–64 years | 784,945 (40.8) |
| 65–74 years | 402,795 (20.9) |
| ≥75 years | 573,181 (29.8) |
| **Region** |  |
| Eastern Japan | 1,106,605 (57.5) |
| Central Japan | 167,134 (8.7) |
| Western Japan | 649,745 (33.8) |

The numbers in parentheses are percentages.

**Table 2.** Volume and cost of low-value care services during April 2022 to March 2023, overall and by age group.

|  | No. of beneficiaries in our sample | Volume |  |  | Spending |  |  |
| --- | --- | --- | --- | --- | --- | --- | --- |
|  |  | Total LVC volume, no. of services | LVC volume per 1,000 beneficiaries | No. (%) of beneficiaries receiving LVC | Total LVC cost, million JPY | LVC cost per 1,000 beneficiaries (million JPY) | % of total healthcare spending within each category <sup>a</sup> |
| **Overall** | 1,923,484 | 3,123,618 | 1623.9 | 696,190 (36.2) | 5329.8 | 2.78 | 0.65 |
| **Stratified by age category** |  |  |  |  |  |  |  |
| <18 years | 162,563 | 228,900 | 1408.1 | 75,219 (46.3) | 79.6 | 0.49 | 0.37 |
| 18–64 years | 784,945 | 538,102 | 685.5 | 196,337 (25.0) | 915.5 | 1.17 | 0.51 |
| 65–74 years | 402,795 | 617,534 | 1533.1 | 140,317 (34.8) | 1183.3 | 2.94 | 0.56 |
| ≥75 years | 573,181 | 1,739,082 | 3034.1 | 284,317 (49.6) | 3151.4 | 5.50 | 0.77 |

<sup>a</sup>Total healthcare spendings in our sample (denominators) were 823.7 billion, 21.5 billion, 180.9 billion, 209.8 billion, and 411.6 billion JPY for the analytic sample overall, aged <18 years, aged 18–64 years, 65–74 years, and ≥75 years.

### Cost Distribution for LVC Volume and Spending

Price categories of very-low-cost, low-cost, medium-cost, and high-cost services consisted of 15, 18, 9, and 10 services, respectively. Among the 3,123,618 LVC services, 3,095,563 were very low cost and low cost (99.1%), compared with only 28,055 such services that were medium cost or high cost (0.9%) (**Figure 1**). The total spending of very-low-cost or low-cost services in our sample (67.3% of unnecessary healthcare spending, or 3.6 billion JPY [28.7 million USD]) exceeded the total spending of medium or high-cost services (32.7% of unnecessary healthcare spending, or 1.7 billion JPY [13.9 million USD]).

**Figure 1.**
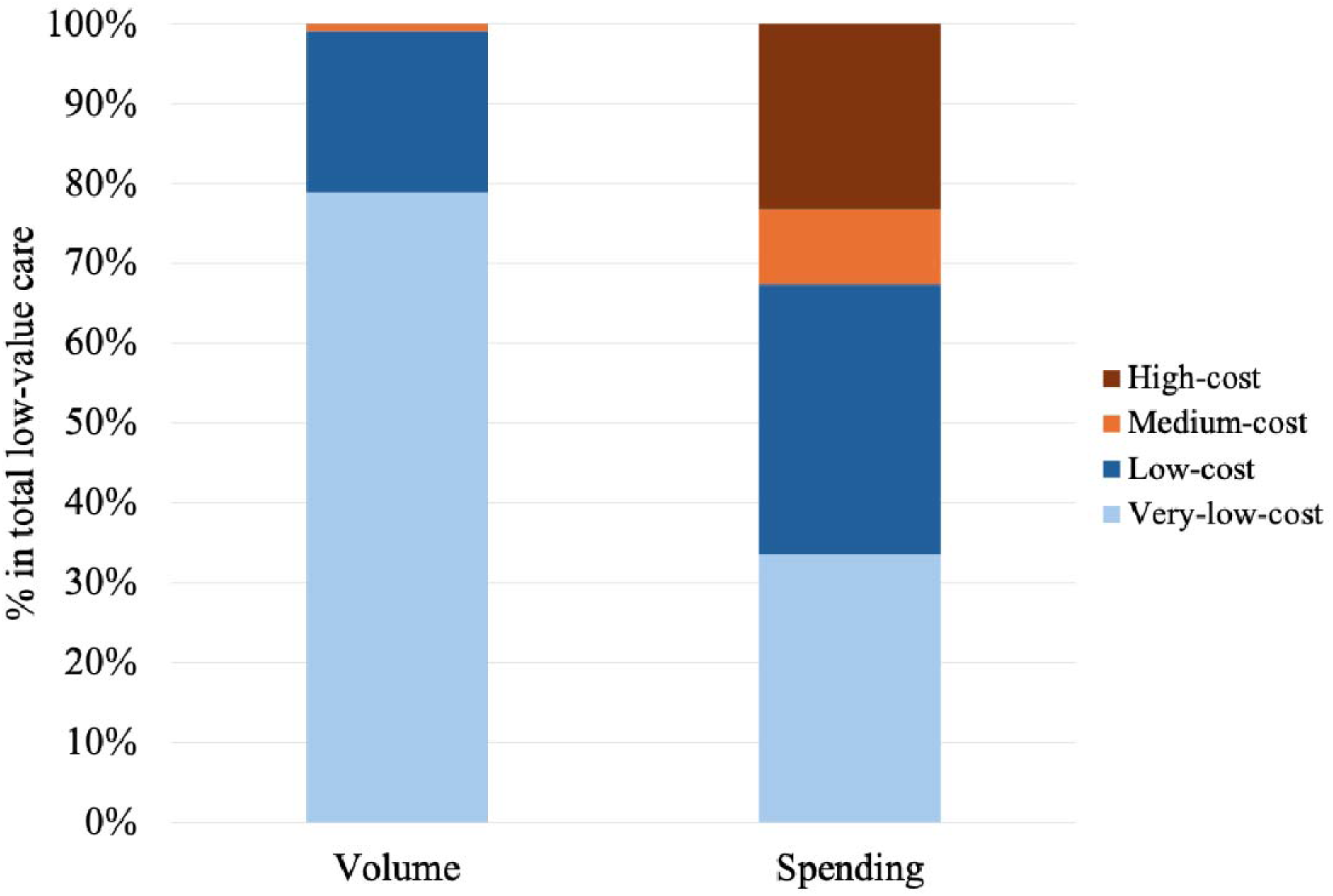
Proportion of total low-value care volume and spending by price category. Low-value care services were classified into four price categories according to average spending per service: very low (< 1,000 Japanese yen [JPY] or 8 US dollars [USD]; 125 JPY = 1 USD in 2022), low (1,000–9,999 JPY [8–80 USD]), medium (10,000–99,999 JPY [80–800 USD]), or high (≥ 100,000 JPY [800 USD]). Categories of very-low-cost, low-cost, medium-cost, and high-cost consisted of 15, 18, 9, and 10 services, respectively.

Five of the top 10 LVC services contributing to unnecessary spending were derived from very-low-cost or low-cost services (**Table 3**). Especially, three services—including topical salicylates or long-term topical nonsteroidal anti-inflammatory drugs (NSAIDs) for chronic pain (very low cost), early imaging for acute low back pain (low cost), and injection for low back pain (low cost)—accounted for approximately half of the grand total LVC spending. All 52 measures are presented in **eTable 4** in **Supplement File**, along with age-, sex-, and region-adjusted national extrapolations.

**Table 3.** The 10 most costly low-value care (LVC) in the total sample.

| **LVC service** | **Price**<br>**(thousand**<br>**JPY) <sup>a</sup>** | **Price**<br>**category <sup>a</sup>** | **Sample**<br>**LVC**<br>**volume** | **Ranking**<br>**by**<br>**volume** | **Sample LVC**<br>**spending (million**<br>**JPY)** | **% in grand total**<br>**LVC spending <sup>b</sup>** |
| --- | --- | --- | --- | --- | --- | --- |
| Topical salicylate or long-term topical nonsteroidal anti-inflammatory drug therapy for chronic pain | 0.9 | Very low | 1,738,631 | 1 | 1532.3 | 28.7 |
| Early imaging for acute low back pain | 3.3 | Low | 211,914 | 3 | 693.7 | 13.0 |
| Vertebroplasty for osteoporotic vertebral fractures | 1463.8 | High | 410 | 31 | 600.2 | 11.3 |
| Injection for low back pain | 6.0 | Low | 74,125 | 9 | 444.0 | 8.3 |
| Imaging for uncomplicated headache | 15.4 | Medium | 19,477 | 13 | 299.9 | 5.6 |
| Percutaneous coronary intervention for stable coronary disease | 1022.6 | High | 242 | 34 | 247.5 | 4.6 |
| Short-term repeat bone mineral testing | 1.5 | Low | 148,452 | 5 | 221.9 | 4.2 |
| Spinal fusion for lumbar stenosis | 243.6 | High | 644 | 28 | 156.9 | 2.9 |
| Pregabalin for low back pain | 1.7 | Low | 84,701 | 8 | 140.6 | 2.6 |
| Preoperative stress testing or stress testing for stable coronary disease | 31.6 | Medium | 4,253 | 20 | 134.6 | 2.5 |

<sup>a</sup>LVC services were classified into four price categories according to average spending per service: very low (< 1,000 Japanese yen [JPY] or 8 US dollars [USD]; 125 JPY = 1 USD in 2022), low (1,000–9,999 JPY [8–80 USD]), medium (10,000–99,999 JPY [80–800 USD]), or high (≥ 100,000 JPY [800 USD]). <sup>b</sup>Grand total LVC cost was the sum of the cost of all 52 low-value care services analyzed in our sample. See **eTable 4** in **Supplement File** for the volume and cost of all 52 low-value care services analyzed.

### Sensitivity Analyses

When using the broader set of LVC definitions (**eTable 5** in **Supplement File**), a total of 3,714,442 LVC services (1931.1 per 1,000 beneficiaries) were provided in 2022-2023, with 39.5% of patients receiving at least one LVC service. The associated spending in this cohort amounted to 8.4 billion JPY (67.2 million USD), representing 1.02% of total healthcare spending. National spending on LVC in Japan, adjusted for age, sex, and region and based on the broader definition, was estimated at 330.5 billion JPY (2.6 billion USD) (**eTable 6** in **Supplement File)**. The findings of the price category distribution were qualitatively unchanged when applying the broader definitions of LVC (**eFigure 1** in **Supplement File**).

The findings of the price category distribution were also qualitatively unchanged by using alternative cutoffs based on the average spending per service (**eFigure 2** in **Supplement File**) and when reanalyzed based on national estimates of LVC volume and spending (**eFigure 3** in **Supplement File**).

### Stratified Analyses by Beneficiaries’ Age

Very-low-cost or low-cost services accounted for most (> 99%) of the volume of LVC services across beneficiaries’ age categories (**Figure 2**). Furthermore, total spending on very-low-cost or low-cost services exceeded that on medium- or high-cost services across all age groups.

**Figure 2.**
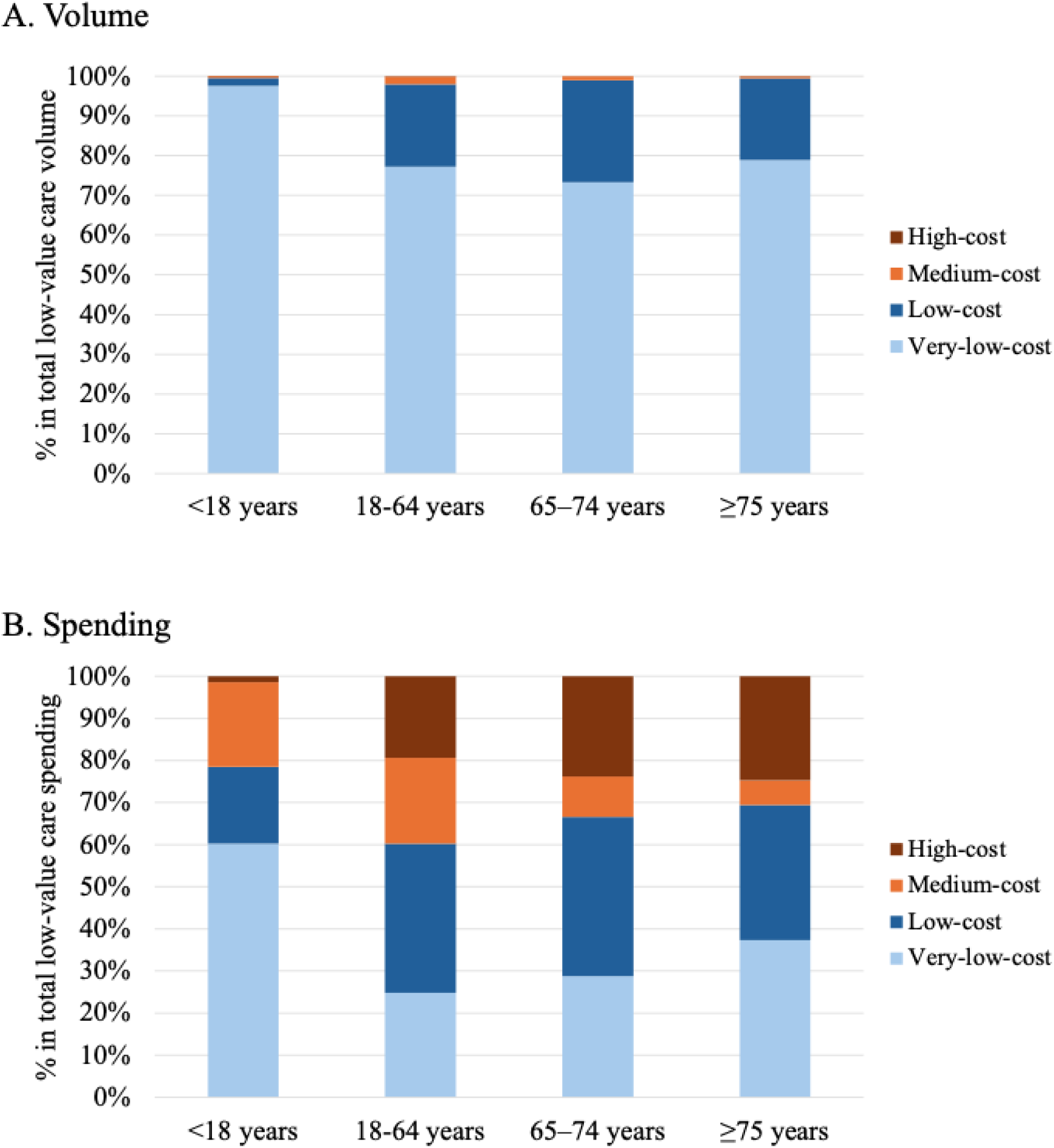
Proportion of total low-value care volume and spending by price category, by beneficiaries’ age group. Low-value care services were classified into four price categories according to an average spending per service: very low (< 1,000 Japanese yen [JPY] or 8 US dollars [USD]; 125 JPY = 1 USD in 2022), low (1,000–9,999 JPY [8–80 USD]), medium (10,000–99,999 JPY [80–800 USD]), or high (≥ 100,000 JPY [800 USD]).

## DISCUSSION

Using a nationwide claims database in Japan, we examined 52 LVC services and found that more than one-third of beneficiaries received at least one LVC service in 2022-2023. The spending for these 52 services alone accounted for 0.7–1.0% of total healthcare spending, corresponding to 207–331 billion JPY (1.7–2.6 billion USD) when extrapolated to the national population with age-, sex-, and region-adjustment. Over 99% of LVC episodes were classified as very-low-cost or low-cost services, and the associated spending on these LVC services exceeded spending for medium-cost or high-cost LVC services. This pattern was consistent across all age groups, including older adults, who often require costly services due to multiple coexisting conditions. These findings suggest that frequently provided low-cost LVC services can cumulatively impose a substantial financial burden on the healthcare system, and represent an important opportunity to identify and reduce wasteful spending.

Interventions to reduce LVC may target policy, medical institutional, physician, and patient levels.^28^ At macro levels, “top-down” approaches to reduce LVC include redesigning supply-side interventions (e.g., payment reforms through global budgets programs^29^ or insurer restrictions^30^) and demand-side strategies (e.g., increasing patient cost-sharing). As suggested by prior literature,^31^ it is important to note that increasing cost-sharing has a higher risk of the unintended consequences of curbing medically necessary care due to patients’ difficulties in choosing care based on value. At micro levels, “bottom-up” approaches, data-driven and non-judgmental dialogue with specialists,^32^ and nudging interventions using electronic health records^33^ have been reported as effective. Enhancing patient education may also be effective.^34^ Ultimately, multi-modal strategies combining these approaches may be most promising.^34^

While high-cost services have frequently been discussed as drivers of rising healthcare spending,^9–13^ our findings suggest that even modest reductions in very-low-cost or low-cost LVC across broad patient populations can cumulatively lead to substantial reduction in unnecessary healthcare spending. Unlike high-cost services, which may have significant financial influence for specific clinical specialties or healthcare sectors, efforts to reduce low-cost LVC are less likely to disrupt provider revenue streams, potentially minimizing policy conflicts.^16^ Nevertheless, designing cost-effective interventions for reducing frequently delivered, low-cost LVC services remains a critical challenge.^35^ For example, broad interventions, such as claims audits targeting all physicians, may not be efficient as de-implementation costs could exceed the low cost of the services. However, prior studies have shown that a large proportion of low-cost LVC is concentrated among a small portion of physicians,^36^ suggesting that targeted interventions focusing on physicians providing high-volume, low-cost services may offer an efficient strategy for maximizing the impact of limited resources.^37^

Among the top 10 LVC services contributing the most to unnecessary spending, low-cost LVC services for pain management—such as topical salicylates or long-term topical NSAIDs therapy for chronic pain, early imaging for acute low back pain, injections for low back pain, and pregabalin for low back pain—were particularly prominent. Collectively, these four services accounted for 52.7% of the total unnecessary spending among the studied measures. Given that low back pain is a leading cause of disability worldwide, including in Japan, reducing LVC in this area is an important public health priority.^38^ The widespread use of LVC services highlights the need to educate physicians on guidelines-based care and to promote high-value and multi-modal alternatives, like exercise programs, physical therapy, yoga, and cognitive behavioral therapy.^39^

Our findings on the price category distribution of unnecessary healthcare spending are consistent with previous studies in the US, where low-cost services contributed more to total LVC spending than high-cost services.^16–18^ The estimated spending due to LVC accounted for 0.65% of total healthcare spending when applying the narrower LVC definition in our study, which is similar to studies in the US that quantified 28 services for the Medicare or commercially insured population.^1,40^ The frequency of LVC (approximately 1.6 per beneficiary per year) in our study was higher than in these US studies. This suggests that Japan’s healthcare system is structurally prone to the provision of low-cost, high-volume LVC. In Japan, hospital-based inpatient care is primarily reimbursed through a bundled payment system, while outpatient services are predominantly reimbursed via a fee-for-service model. This system can create incentives for physicians to frequently provide low-cost LVC in outpatient settings. This tendency may be influenced by the characteristics of Japan’s health system, including low patient cost-sharing (particularly for those aged 75 and older, who pay only 10% unless they are high-income earners) and unrestricted access to clinics and hospitals.

Our study has limitations. First, as with any study measuring LVC directly,^1,7,16,27^ our analysis was subject to the inherent limitations of using administrative claims data. Although claims data reliably indicate whether a given procedure was performed, they often lack the detailed clinical context required to determine the appropriateness of that care—information more commonly available in medical records. To address this limitation, we selected definitions of LVC with higher specificity to better capture instances of overuse. Despite these constraints, claims data represent a cost-efficient alternative to medical record review and are particularly valuable for continuous monitoring and the development of payment policies.^1^ Second, although we included 52 LVC services, our measures were limited to those assessable using claims data. For example, because symptoms such as frequent or painful urination cannot be reliably identified in claims data, we could not evaluate bacteriuria screening in asymptomatic patients, which reportedly accounts for about 0.1% (1.2 billion USD) of total Medicare spending in the US.^41^ Furthermore, preventive services, like prostate-specific antigen screening for older men, were not covered by public insurance in Japan and thus not captured. However, including these would likely not change our cost distribution findings, as such services are typically low-cost despite frequent use. Third, we did not capture downstream costs associated with LVC, such as healthcare spending on adverse events and complications associated with LVC,^42–45^ which may have resulted in an underestimation of total unnecessary spending. For example, a recent study demonstrated that when indirect costs from downstream testing and treatments triggered by low-value preoperative electrocardiograms before cataract surgery are included, the associated costs increase by a factor of ten.^43^ Despite these limitations, our estimates indicate a substantial burden of LVC services in Japan. Finally, the claims data we used were based on a convenient sample, leading to a slightly older study population compared to the general Japanese population. However, findings were qualitatively unchanged when stratified by beneficiaries’ age or when using age-sex-region national extrapolations, suggesting generalizability to the entire Japanese population.

## CONCLUSION

This nationwide study in Japan found that low-cost, high-volume services contributed substantially to unnecessary healthcare spending across all age groups. Rather than solely targeting high-cost services, curbing the overuse of frequently performed, lower-cost services may offer a more effective approach to reducing LVC.

## Data Availability

The DeSC database is a proprietary database owned by DeSC Healthcare, Inc.; therefore, it cannot be shared. Individuals who are interested in using the DeSC data should contact DeSC Healthcare, Inc.

https://desc-hc.co.jp/en

## Acknowledgments

The authors would like to acknowledge the contribution of the following team of specialist physicians who reviewed clinical evidence and developed a candidate list of low-value care services. The team members are as follows: Dr. Yuichiro Matsuo; Dr. Junya Arai; Dr. Naoko Hidaka; Dr. Hideaki Watanabe; Dr. Yuki Uehara; Dr. Masashi Miyawaki; Dr. Arisa Iba; Dr. Shinichiro Shiomi; Dr. Yu Mizushima; Dr. Teiichiro Aoyagi; Dr. Yoshio Hayashi; Dr. Asahi Fujita; Dr. Risa Ishida; Dr. Takuya Sato; Dr. Ayako Yanagisawa; and Dr. Shinji Sugahara.

## Author Contributions

Dr. Miyawaki had full access to the study data and takes responsibility for the accuracy and integrity of the data and analyses. Study concept and design: Miyawaki, Tsugawa. Acquisition, analysis, or interpretation of data: All authors. Drafting of the manuscript: Miyawaki. Critical revision of the manuscript for important intellectual content: All authors. Statistical analysis: Miyawaki. Administrative, technical, or material support: All authors.

## Role of the Funder or Sponsor

None.

## Data-Sharing Statement

The DeSC database is a proprietary database owned by DeSC Healthcare, Inc.; therefore, it cannot be shared. Individuals who are interested in using the DeSC data should contact DeSC Healthcare, Inc. (https://desc-hc.co.jp/en).

