## Supplement File for "Low-Cost, High-Volume Health Services Contribute the Most to Unnecessary Health Spending due to Low-Value Care in Japan"

**eMethod 1. Low-value care identification**

This study examined 52 low-value care (LVC) services based on a comprehensive approach that combined our previous study and a detailed literature review. Initially, we included 31 LVC services, excluding two services that were not provided in that study (unblocking nasolacrimal duct in infants <12 months and electroconvulsive therapy in children with depression) among the 33 LVC services identified in previous research conducted in Japan.^1^ To ensure a more thorough assessment using a comprehensive set of LVC services in Japan, we further conducted a pre-determined literature review process to expand the set of LVC services. This process was based on the candidates of LVC selected by an expert group, following the methodology used in the previous study.

First, we began by assembling a specialist physician board covering as many clinical specialty areas as possible. The specialists were selected by the author group based on experience in their specialty, sex, and geographical location. The selected specialists reviewed clinical evidence from peer-reviewed medical literature to compile a list of potentially LVC services in their respective specialty areas, allowing for overlap. Additionally, based on the experts’ assessment, the level of evidence for each service was categorized into the following three groups: (1) Definitely low-value (there is evidence that it has no clinical benefit, namely the service has been concluded to be “having no effect” in meta-analyses or multiple studies including randomized controlled trials, with minimal variation in results), (2) Possibly low-value (meta-analyses or multiple studies including randomized controlled trials show no effect, but with large variation in results [some groups benefit]), and (3) Unclear (there is no evidence that it has clinical benefit). We contacted physicians in 23 specialties and received responses from physicians in 19 specialties (general internal medicine, family medicine, respiratory medicine, gastroenterology, diabetes, endocrinology, nephrology, infectious disease medicine, geriatrics, rheumatology, general surgery, otolaryngology, urology, dermatology, ophthalmology, obstetrics and gynecology, emergency medicine, anesthesiology, and radiology), excluding the four specialties of hematology, cardiac surgery, psychiatry, and neurosurgery. The specialists identified a total of 138 candidate services of LVC.

Second, for each candidate service of LVC listed by the specialists, independent physicians (A.M. and Y.K. from the author group) repeated the literature review and categorization process using the same methodology. Services consistently categorized as “definitely low value” by both specialists and independent physicians were selected as LVC. Among the 140 candidate services, 44 services met this criterion.

Third, we selected LVC services measurable in Japanese hospital claims data. The determination of measurability was conducted by two physicians with substantial experience in claims data analyses (A.M. and Y.K.). The selection criteria were: (1) The service is recorded in hospital claims data under the Japanese health insurance system, and (2) The service can be reasonably identified as a low-value episode using the available variables with high specificity. Services deemed unmeasurable included those lacking sufficient information in the claims data for identification (e.g., bacteriuria screening for asymptomatic patients) and those not covered by Japanese public health insurance (preventive care services, prostate cancer screening using prostate-specific antigen [PSA] testing at age >75 years, and pregnancy checkups are not covered). We excluded 20 unmeasurable LVC items (details are shown in **eTable 3** below), resulting in 24 measurable LVC services.

Finally, we integrated these 24 LVC items from our literature review with the 31 LVC items identified in our previous study. After removing three duplicates, we identified 52 LVC items.

**eTable 1** shows each LVC service measure and its operational definition. As was the case with previous studies,^19,33^ when there were both sensitive (broader) and specific (narrower) definitions for defining metrics, we used the more specific definition. This approach reduces the likelihood of detecting LVC services but also reduces the likelihood of misclassifying high-value services as LVC services. We used the operational definitions established by the consensus method in previous research conducted in Japan^27^ with some modifications for application to the DeSC claims data. For the 21 newly added LVC services, definitions were established through a consensus method by three physicians experienced in quantifying services in healthcare administrative data (A.M., Y.K., Y.T.), following the approach used in previous research. Codes for measures of LVC services were shown in **eTable 2.**

**eMethod 2. Age-, Sex-, and Region-adjusted national extrapolations**

Our sample was a convenience sample. Because the database was not nationally representative, we estimated the national volume and spending on low-value care (LVC) by extrapolating our findings to the national population with adjustment for age, sex, and region.

For each LVC measure, we calculated per capita volume and spending within strata defined by five-year age groups (0–4, 5–9, …, 80–84, and ≥85 years), sex (male/female), and region (Eastern, Central, or Western Japan), resulting in 18 × 2 × 3 = 108 strata. We then multiplied these stratum-specific values by the corresponding 2022 population counts and summed across all strata to obtain national estimates. This method accounted for demographic and geographic variation in healthcare use and provided more policy-relevant national estimates.

**eTable 1. Operational definitions of low-value care (LVC) for each healthcare service**

| **Healthcare service** | **Service denominator** | **Numerator** | |
| --- | --- | --- | --- |
|  |  | **Broader LVC definition**  **(Base definition)** | **Narrower LVC definition**  **(Additional restrictions)** |
| **Drugs (N=21)** |  |  |  |
| Antibiotics prescription for acute pancreatitis ^2,3^ | Any antibiotics prescription (oral/intravenous) other than imipenem | Antibiotics prescription of other than imipenem within three days of admission for hospitalized patients with an admission diagnosis of acute pancreatitis | No diagnosis of biliary acute pancreatitis and cholangitis |
| Antibiotics prescription for acute respiratory infections (ARIs) ^4^ | Any oral antibiotics prescription | Oral antibiotics prescription for patients with a diagnosis of ARIs (prescriptions on the day of diagnosis) | No diagnoses for which antibiotics may be appropriate^a^ |
| Antibiotics prescription for threatened preterm labor ^5^ | Any antibiotics prescription (oral/intravenous) | Antibiotics prescription for women with a diagnosis of threatened preterm labor (prescriptions on the day of diagnosis) | No diagnosis of other obstetric complications necessitating antibiotics ^b^ |
| Antibiotics prescription for uncomplicated colon diverticulitis ^6^ | Any outpatient antibiotics prescription (oral/intravenous) | Antibiotics prescription for patients with a diagnosis of uncomplicated colon diverticulitis (prescriptions on the day of diagnosis) | No diagnosis of complicated colon diverticulitis |
| Antihistamine-analgesic combinations for ARIs ^7^ | Any oral antihistamine-analgesic combination prescription | Antihistamine-analgesic combination prescriptions for patients with a diagnosis of ARIs (prescriptions on the day of diagnosis) | No diagnoses for which antibiotics may be appropriate ^a^ |
| Antihistamines for ARIs ^8^ | Any oral Antihistamine prescription | Antihistamine prescriptions (excluding antihistamine-analgesic combinations) for patients with a diagnosis of ARIs (prescriptions on the day of diagnosis) | No diagnoses for which antibiotics may be appropriate ^a^ or allergy diseases |
| Codeines prescription for ARIs ^9,10^ | Any oral codeines prescription | Codeines prescription for patients with a diagnosis of ARIs (prescriptions on the day of diagnosis) | No diagnosis for which antibiotics may be appropriate,^a^ chronic respiratory diseases, or chronic pain |
| Dextromethorphan prescription for ARIs in children ^11–13^ | Any oral dextromethorphan prescription for children aged < 18 years | Dextromethorphan prescription for children with a diagnosis of ARIs (prescriptions on the day of diagnosis) | No diagnoses for which antibiotics may be appropriate ^a^ or chronic respiratory diseases |
| Expectorants prescription for ARIs ^14^ | Any oral expectorants prescription | Expectorants prescription for patients with a diagnosis of ARIs (prescriptions on the day of diagnosis) | No diagnoses for which antibiotics may be appropriate ^a^ or chronic respiratory diseases |
| Intravenous anti-herpes drugs for sudden sensorineural hearing loss ^15^ | Any hospitalizations with intravenous acyclovir administration | Hospitalizations with intravenous anti-herpes drugs administration for a patient with a diagnosis of sudden sensorineural hearing loss | Only patients without a diagnosis of herpes infections (herpes simplex virus or herpes zoster virus) |
| Intravenous betamimetics for inhibiting preterm labor, > 48h ^16^ | Any hospitalizations with intravenous betamimetics administration | Hospitalizations in which betamimetics intravenous infusion was administered for ≥ 3 days | The narrower definition is set the same as the broader definition. |
| Intravenous sivelestat for acute respiratory disease syndrome ^17,18^ | Any hospitalizations with intravenous sivelestat administration | Any hospitalizations with intravenous sivelestat administration | The narrower definition is set the same as the broader definition. |
| Neuraminidase inhibitors for non-severe influenza ^19^ | Any neuraminidase inhibitor prescriptions in adults aged ≥ 18 years | Any oseltamivir, laninamivir, or zanamivir prescriptions for adult patients with influenza (prescriptions on the day of diagnosis) | No diagnoses for which antibiotics may be appropriate^a^ or influenza pneumonia/encephalopathy |
| Nitroglycerin or isosorbide dinitrate for perioperative management of non-cardiac surgery ^20^ | Intravenous nitroglycerin or isosorbide dinitrate administration | Nitroglycerin or isosorbide dinitrate for patients undergoing a low or intermediate-risk non-cardiothoracic surgical procedures (selected surgeries) from one day before to 30 days after the surgery | No diagnosis of acute coronary syndrome |
| Oral betamimetics prescription ^21^ | Any oral betamimetics prescription | Any oral betamimetics prescription | The narrower definition is set the same as the broader definition. |
| Pregabalin prescription for back pain ^22,23^ | Any pregabalin prescription | Pregabalin prescription in a patient with diagnosis of back pain | Pregabalin prescription in a patient with diagnosis of back pain (except for patients with a diagnosis of fibromyalgia, diabetes, postherpetic neuralgia, arteriosclerosis, disk disorder, trigeminal neuralgia, or peripheral neuropathy) |
| Topical salicylates or long-term topical non-steroidal anti-inflammatory drugs (NSAIDs) therapy for chronic pain ^24,25^ | Any topical salicylates or topical NSAIDS prescription | 1) Any salicylates prescription  2) Topical NSAIDs prescriptions exceeding three months in the year | The narrower definition is set the same as the broader definition. |
| Tranexamic acid administration for intracerebral hemorrhage ^26^ | Any hospitalizations with intravenous tranexamic acid administration | Hospitalizations with tranexamic acid administration for patients with an admission diagnosis of intracerebral hemorrhage | The narrower definition is set the same as the broader definition. |
| Tricyclic antidepressants prescription for children without other psychological disorders ^27^ | Any tricyclic antidepressants prescription for children in children aged 6-18 years | Tricyclic antidepressants prescription for children with a diagnosis of depression | Tricyclic antidepressants prescription for children with a diagnosis of depression, and without other psychological disorders |
| Ursodeoxycholic acid prescription for primary biliary cirrhosis, alcoholic cirrhosis, primary sclerosing cholangitis ^28^ | Any oral ursodeoxycholic acid prescription | Oral ursodeoxycholic acid prescription for patients with a diagnosis of primary biliary cirrhosis, alcoholic liver disease with jaundice, primary sclerosing cholangitis | No diagnosis of cholelithiasis |
| Vitamin B12 medications for diabetic neuropathy ^29^ | Any oral vitamin B12 medication prescription | Vitamin B12 medication prescription for a patient with a diagnosis of diabetes mellitus, excluding vitamin B12 deficiency-related conditions | Only patients with a diagnosis of diabetic neuropathy |
| **Laboratory or imaging tests (N=14)** |  |  |  |
| 1, 25-dihydroxy vitamin D testing in the absence of hypercalcemia or decreased kidney function ^30^ | Any calcitriol testing in adults aged ≥ 18 years | Calcitriol testing for patients without hypercalcemia or secondary hyperparathyroidism was noted in the claim, and without a history of CKD | No diagnoses indicating non-PTH mediated hypercalcemia (sarcoidosis, tuberculosis, and selected neoplasms) |
| Bone mineral density testing at frequent intervals ^31–33^ | Any bone mineral density test | Subsequent bone mineral density test in the year | Only patients with a diagnosis of osteoporosis at the timing of the initial bone mineral density test |
| Early imaging for acute low back pain ^34,35^ | Any plain x-ray, computed tomography (CT), or magnetic resonance imaging (MRI) | Plain x-ray, CT, or MRI conducted on the date of low back pain diagnosis ^c^ | No diagnoses in claim warranting imaging |
| Electroencephalography (EEG) for headache ^36^ | Any outpatient EEG | EEG with headache diagnosis in the claim | No epilepsy or convulsions in the claims |
| Frequent pulmonary function test (PFT) for stable chronic obstructive pulmonary disease (COPD) ^37^ | Any outpatient PFT | Second or subsequent PFT in a year for patients with a diagnosis of COPD, not occurring within 30 days prior to a low or intermediate-risk surgical procedure (selected surgeries) (to make mutually exclusive for preoperative PFT) and without any hospital admissions within the past year | No diagnosis of chronic pulmonary diseases (except for COPD) |
| Hypercoagulability testing for patients with deep vein thrombosis ^38^ | Any lab tests for hypercoagulable states | Laboratory tests for hypercoagulable states in a patient with a diagnosis of lower extremity deep vein thrombosis or pulmonary embolism | The narrower definition is set the same as the broader definition. |
| Imaging for uncomplicated headache ^39–42^ | Any CT or MRI | CT or MRI conducted on the date of low headache diagnosis  CT or MRI with nonposttraumatic, nonthunderclap headache (imaging occurring on the day of diagnosis) | No diagnoses in claim warranting imaging |
| PTH testing for patients with stage 1-3 chronic kidney disease ^43,44^ | Any PTH testing | PTH measurement in a patient with chronic kidney disease, without dialysis services in any of the same fiscal year claims | No hypercalcemia diagnosis in any of the same fiscal year claims |
| Preoperative breast MRI ^45^ | Any breast MRI in adults aged ≥ 18 | Breast MRI within 30 days prior to breast cancer surgery | Only patients with no diagnosis of hereditary breast and ovarian cancer syndrome and breast Paget's disease, no surgery for benign breast diseases within three months, and no diagnosis of breast cancer more than 12 months prior to the MRI (to include only primary breast cancer) |
| Preoperative echocardiogram ^46^ | Any outpatient echocardiograms in adults aged ≥ 18 years | Echocardiogram occurring within 30 days prior to a low- or intermediate-risk non-cardiothoracic surgical procedure (selected surgeries) | No echocardiograms related to off-hour visits |
| Preoperative PFT ^47^ | Any outpatient PFT in adults aged ≥ 18 years | PFT occurring within 30 days prior to a low or intermediate-risk surgical procedure (selected surgeries) | No PFTs related to off-hour visits |
| Preoperative stress testing or stress testing for stable coronary disease ^48–50^ | Any outpatient stress electrocardiogram, echocardiogram, nuclear medicine imaging, cardiac MRI, or CT angiography in adults aged ≥ 18 years | 1) Stress test occurring within 30 days prior to a low- or intermediate-risk non-cardiothoracic surgical procedure (selected surgeries) or 2) stress test in a patient with a diagnosis of chronic ischemic heart disease | No stress test related to off-hour visits, which might be indicative of acute coronary syndrome |
| Routine preoperative medical testing | Routine preoperative medical testing (blood test, x-ray, and electrocardiogram) | Medical testing within 30 days prior to a low- or intermediate-risk non-cardiothoracic surgical procedure | Medical testing for patients undergoing cataract surgery within 30 days prior to the surgery; only preoperative medical testing not related to off-hour visits within 30 days prior to the surgery |
| Serum T3 level testing for patients with hypothyroidism ^30^ | Any total or free T3 test | Total or free T3 measurement in a patient with a hypothyroidism diagnosis during that calendar year | The narrower definition is set the same as the broader definition. |
| **Procedures (N=17)** |  |  |  |
| Arthroscopic surgery for knee osteoarthritis ^55,56^ | Any arthroscopic debridement/chondro-plasty of the knee in adults aged ≥ 18 years | Arthroscopic surgery of the knee with the diagnosis of osteoarthritis or chondromalacia in the claim | No meniscal tear noted in the claim |
| Artificial liver support for acute liver failure ^57^ | Any hospitalizations with artificial liver support in adults aged ≥ 18 years | Hospitalizations with artificial liver support in a patient with a diagnosis of acute liver failure | Only hospitalizations without a diagnosis of chronic or subacute liver disease |
| Bed rest for threatened preterm labor ^58^ | Any hospitalization for women with diagnoses of pregnancy-related conditions | Hospitalization of patients with an admission diagnosis of threatened preterm labor, not using tocolytic drugs and systemic corticosteroids | No diagnosis of any other obstetric complications necessitating hospitalization^b^ |
| Carotid endarterectomy in asymptomatic patients ^59,60^ | Any carotid endarterectomy in adults aged ≥ 18 years | Carotid endarterectomy for a patient aged ≥ 75 without a diagnosis of stroke, transient ischemic attack TIA, retinal artery occlusion, or nervous and musculoskeletal symptoms. Exclude emergency admissions (identified in inpatient claims) | Only a patient with a history of acute myocardial infarction, chronic obstructive pulmonary disease, or alcohol related disorder, with dialysis services, with presence of cardiac pacemaker, or aged 90 or older (indicating high-risk patients) |
| Emergency surgery for acute appendicitis shortly after admission (<24 h) (literature review) ^51–53^ | Surgery for acute appendicitis | Emergency surgery for a patient with a diagnosis of acute appendicitis without peritonitis shortly after admission | No diagnosis of peritonitis |
| Endoscopy for dyspepsia for people < 55 years or colonoscopy for constipation in people < 50 years ^61–65^ | Any endoscopies (esophagogastroduo-denoscopies and colonoscopies) in adults aged 18-54 | 1) Endoscopy in a patient aged 18-54 with a diagnosis of dyspepsia or 2) colonoscopy in a patient aged 18-49 with a diagnosis of constipation | 1) Endoscopy in a person aged 18-54 with a diagnosis of dyspepsia and no diagnoses of dysphagia, anemia, weight loss, cancer of the digestive system or 2) colonoscopy in a person aged 18-49 with a diagnosis of constipation and no diagnoses of anemia, weight loss, cancer of the digestive system, or other diseases of the digestive system |
| Endotoxin apheresis for sepsis ^66,67^ | Any hospitalizations with endotoxin apheresis in adults aged ≥ 18 years | Hospitalizations with endotoxin apheresis | The narrower definition is set the same as the broader definition. |
| Feeding tube for severe dementia ^68^ | Feeding tube | Gastrostomy, percutaneous trans-esophageal gastro-tube insertion, elemental diet tube insertion, and tube feeding management for patients with dementia who are receiving home healthcare or institutional care | No diagnosis of Crohn’s disease, ulcerative colitis, ulcer of the esophagus, esophageal obstruction, perforation of esophagus, head injury and burn, congenital malformations of esophagus, or diseases of nervous system other than dementia |
| Inferior vena cava (IVC) filters for the prevention of pulmonary embolism ^69,70^ | Any hospitalizations with IVC filter placement in adults aged ≥ 18 years | Hospitalizations with IVC filter placement in a patient with a diagnosis of deep vein thrombosis or pulmonary embolism | The narrower definition is set the same as the broader definition. |
| Percutaneous coronary intervention (PCI) with angioplasty or stent placement for stable coronary disease ^50,71^ | Any stent placement or balloon angioplasty in adults aged ≥ 18 years | Stent placement or balloon angioplasty for a patient with an established diagnosis of ischemic heart disease or angina (at least six months prior to the procedure), not related to acute coronary syndrome and not associated with off-hour visit, which might be indicative of acute coronary syndrome | Only patients with a past diagnosis of myocardial infarction in order to exclude patients with a history of non-cardiac chest pain inaccurately coded as angina |
| Pulmonary artery catheterization (PAC) in the ICU ^72^ | Any hospitalizations with PAC in adults aged ≥ 18 years | PAC during an inpatient stay that involved an ICU but not surgical procedures | Exclude claims that involved pulmonary hypertension or cardiac tamponade |
| Renal angioplasty ^73,74^ | Any renal angioplasty in adults aged ≥ 18 years | Any renal angioplasty | Only a patient with a diagnosis of renovascular hypertension or renal atherosclerosis, and no diagnosis of fibromuscular dysplasia of renal artery, in the claim |
| Spinal fusion for lumbar stenosis ^75,76^ | Any hospitalizations with spinal fusion surgeries in adults aged ≥ 18 | Spinal fusion surgeries for a patient with a diagnosis of lumbar stenosis | Only patients without a diagnosis of spondylolisthesis, pain in the foot, radiculopathy, sciatica, or congenital malformations of the spine |
| Spinal injection for low back pain ^77,78^ | Any outpatient spinal injection in adults aged ≥ 18 years | Epidural (not indwelling), facet, or trigger point injections in a patient with a diagnosis of lower back pain in the same month, not associated with an inpatient stay (within 14 days) | No diagnoses indicating radiculopathy in the claim in the same month |
| Surgery for vesicoureteral reflux ^79,80^ | Any surgery for vesicoureteral reflux in children aged <12 years | Any surgery for a patient with a diagnosis of vesicoureteral reflux | Only patients with vesicoureteral reflux |
| Traction therapy for back pain or neck pain ^81–83^ | Any traction therapy in adults aged ≥ 18 years | Traction therapy for a patient with a diagnosis related to back or neck pain | Only patients without a diagnosis of malignant spinal tumor, rheumatoid arthritis, spondylitis, or osteoporosis |
| Vertebroplasty for osteoporotic vertebral fractures ^84–86^ | Any vertebroplasty in adults aged ≥ 18 years | Vertebroplasty for a patient with a diagnosis of vertebral fracture | No bone cancers, myeloma, or hemangioma were noted in the claim in the same month |

^a^ Conditions for which antibiotics may be appropriate include acute sinusitis, acute pharyngitis, tonsillitis, acute tracheitis, acute epiglottitis, acute bacterial pneumonia, unspecified acute lower respiratory infection, peritonsillar abscess, chronic pharyngitis, chronic sinusitis, otitis media.

^b^ Other obstetric complications necessitating hospitalization include premature rupture of membranes, eclampsia, HELLP syndrome, multiple pregnancy, hypertensive disorders of pregnancy, diabetes, antepartum hemorrhage, other disorders of amniotic fluid and membranes, placental abruption, and placenta previa. Other obstetric complications necessitating antibiotics include infections of genitourinary tract in pregnancy, infection of amniotic sac and membranes (chorioamnionitis), and premature rupture of membranes.

^c^ Although back imaging within six weeks after symptom onset is not recommended, the Japanese claims data include imaging procedure codes without detailed information on the body region imaged, which makes it challenging to precisely identify back imaging. Given that imaging performed on the date of a low back pain diagnosis can reasonably be assumed to be back imaging, we limited our analysis to imaging procedures conducted on the diagnosis date.

**eTable 2. Codes for measures of low-value care services**

| **Healthcare services** | **Codes for identification** | **Healthcare spending calculation** |
| --- | --- | --- |
| **Drugs (N=21)** |  |  |
| Antibiotics prescription for acute pancreatitis | **Anatomical Therapeutic Chemical (ATC) classification system code:** J01 (any antibiotics); J01DH51 (imipenem). **International Classification of Diseases, Tenth Revision (ICD-10) code:** K85 (acute pancreatitis); K851 (biliary acute pancreatitis); K830 (cholangitis) | a |
| Antibiotics prescription for acute respiratory infections (ARIs) | **ATC code:** J01 (any antibiotics) **ICD-10 code:** J00-J06 (ARIs); J01 (acute sinusitis); J020 (acute pharyngitis); J030 (tonsillitis); J041 (acute tracheitis); J051 (acute epiglottitis); J13-J18 (acute bacterial pneumonia); J22 (unspecified acute lower respiratory infection); J36 (peritonsillar abscess); J312 (chronic pharyngitis); J32 (chronic sinusitis); H66 (otitis media) | a |
| Antibiotics prescription for threatened preterm labor | **ATC code:** J01 (any antibiotics) **ICD-10 code:** O600 (threatened preterm labor); O23 (infections of genitourinary tract in pregnancy); O411 (infection of amniotic sac and membranes (chorioamnionitis)); O42 (premature rupture of membranes) | a |
| Antibiotics prescription for uncomplicated colon diverticulitis | **ATC code:** J01 (any antibiotics) **ICD-10 code:** K573, K575, K579 (uncomplicated colon diverticulitis); K570, K572, K574, K578 (complicated intestine diverticulitis) | a |
| Antihistamine-analgesic combinations for ARIs | **ATC code:** R05X **ICD-10 code:** J00-J06 (ARIs); J01 (acute sinusitis); J020 (acute pharyngitis); J030 (tonsillitis); J041 (acute tracheitis); J051 (acute epiglottitis); J13-J18 (acute bacterial pneumonia); J22 (unspecified acute lower respiratory infection); J36 (peritonsillar abscess); J312 (chronic pharyngitis); J32 (chronic sinusitis); H66 (otitis media) | a |
| Antihistamines for ARIs | **ATC code:** R06 **ICD-10 code:** J00-J06 (ARIs); J01 (acute sinusitis); J020 (acute pharyngitis); J030 (tonsillitis); J041 (acute tracheitis); J051 (acute epiglottitis); J13-J18 (acute bacterial pneumonia); J22 (unspecified acute lower respiratory infection); J36 (peritonsillar abscess); J312 (chronic pharyngitis); J32 (chronic sinusitis); H66 (otitis media); H011, H101, J30, L20, L29, L50 (allergy diseases) | a |
| Codeine prescription for the ARIs | **ATC code**: N02AA08, N02AA58, R05DA04, R05DA12 (codeines) **ICD-10 code:**ICD-10: J00-J06 (ARIs); J01 (acute sinusitis); J020 (acute pharyngitis); J030 (tonsillitis); J041 (acute tracheitis); J051 (acute epiglottitis); J13-J18 (acute bacterial pneumonia); J22 (unspecified acute lower respiratory infection); J36 (peritonsillar abscess); J312 (chronic pharyngitis); J32 (chronic sinusitis); H66 (otitis media); J41, J42, J43, J44, J45, J47, J6x, J84, C33, C34 (chronic respiratory diseases); R521, R522 (chronic pain) | a |
| Dextromethorphan prescription for ARIs in children | **ATC code:** R05DA09 (dextromethorphan) **ICD-10 code:**ICD-10: J00-J06 (ARIs); J01 (acute sinusitis); J020 (acute pharyngitis); J030 (tonsillitis); J041 (acute tracheitis); J051 (acute epiglottitis); J13-J18 (acute bacterial pneumonia); J22 (unspecified acute lower respiratory infection); J36 (peritonsillar abscess); J312 (chronic pharyngitis); J32 (chronic sinusitis); H66 (otitis media); J41, J42, J43, J44, J45, J47, J6x, J84, C33, C34 (chronic respiratory diseases) | a |
| Expectorant prescription for ARIs | **ATC code:** R05CB01, R05CB03 (expectorants). **ICD-10 code:** J00-J06 (ARIs); J01 (acute sinusitis); J020 (acute pharyngitis); J030 (tonsillitis); J041 (acute tracheitis); J051 (acute epiglottitis); J13-J18 (acute bacterial pneumonia); J22 (unspecified acute lower respiratory infection); J36 (peritonsillar abscess); J312 (chronic pharyngitis); J32 (chronic sinusitis); H66 (otitis media); J41, J42, J43, J44, J45, J47, J6x, J84, C33, C34 (chronic respiratory diseases) | a |
| Intravenous anti-herpes drugs for sudden sensorineural hearing loss | **Japanese Medical Practice Code:** 620001341, 620003671, 620003746, 620004633, 620006283, 620006284, 620009268, 621144901, 621384302, 621384303, 621384402, 621384411, 621384414, 621384422, 621384424, 621384425, 621660102, 622325900, 640461002 (intravenous acyclovir)  **ICD-10 code:** H912 (sudden sensorineural hearing loss); A60 (herpes simplex infection), B00-B02 (herpes zoster virus infection); B203 (human immunodeficiency virus diseases in herpes virus infections) | a |
| Intravenous betamimetics for inhibiting preterm labor, >48h | **Japanese Medical Practice Code:** G02CA01 (betamimetics, intravenous rout) | a |
| Intravenous sivelestat for acute respiratory disease syndrome | **Japanese Medical Practice Code:** 622381901, 622391901, 622393901, 622394601, 622401101, 640462009 (sivelestat) | a |
| Neuraminidase inhibitors for non-severe influenza | **ATC code:** J05AH02 (oseltamivir); J05AH04 (laninamivir); J05AH01 (zanamivir).  **ICD-10 code:** J09, J10, J11 (influenza); J01 (acute sinusitis); J020 (acute pharyngitis); J030 (tonsillitis); J041 (acute tracheitis); J051 (acute epiglottitis); J13-J18 (acute bacterial pneumonia); J22 (unspecified acute lower respiratory infection); J36 (peritonsillar abscess); J312 (chronic pharyngitis); J32 (chronic sinusitis); H66 (otitis media); H011, H101, J30, L20, L29, L50 (allergy diseases); J100, J108, J110, J118 (Influenza pneumonia/encephalopathy) | a |
| Nitroglycerin or isosorbide dinitrate for perioperative management of non-cardiac surgery | **ATC code:** C01DA02 (nitroglycerin); C01DA08 (isosorbide dinitrate)  **ICD-10 code:** I20, I21, I22, I24 (acute coronary syndrome).  **Billing Category code ^d^:** Low- or intermediate-risk non-cardiothoracic surgical procedures include surgeries of the breast (K472-K476), colectomy (K719), cholecystectomy (K672), transurethral resection of the prostate (K841), hysterectomy (K872-K879), orthopedic surgeries including arthroscopy (besides hip and knee replacement) (K023-K144), corneal transplant (K259), cataract removal (K282), retinal detachment (K275, K276, K277, K279, K280, K281, K284), hernia repair (K633, K634), and lithotripsy (K768) | a |
| Oral betamimetics prescription | **ATC code:** G02CA01 (oral betamimetics) | a |
| Pregabalin prescription for back pain | **ATC code:** N02BF02 (pregabalin)  **ICD-10 code:** M4306, M4309, M4316, M4317, M4319, M4329, M4646, M4649, M4716, M4719, M4786, M4789, M4799, M4800, M4806, M4808, M4809, M510, M511, M512, M513, M518, M519, M5326, M5328, M5329, M533, M539, M5416–M5419, M543, M544, M545, M548, M549, M961, M99, Q762, S33 (back pain); M797 (fibromyalgia); B022, G530 (postherpetic neuralgia); M50, M51 (disk disorder); G50 (trigeminal neuralgia); E10-E14 (diabetes); I70 (arteriosclerosis); T812, G54-G64 (peripheral neuropathy) | a |
| Topical salicylates or long-term topical nonsteroidal anti-inflammatory drugs (NSAIDs) therapy for chronic pain | **ATC code:** M02AC (Topical salicylates); M02AA (topical NSAIDS) | a |
| Tranexamic acid administration for intracerebral hemorrhage | **ATC code:** B02AA02 (tranexamic acid) **ICD-10 code:** I61 (intracerebral hemorrhage) | a |
| Tricyclic antidepressants prescription for children without other psychological disorders | **ATC code:** N06AA (oral tricyclic antidepressants)  **ICD-10 code:** F32, F33 (depression); F01-F31, F34-F99 (other psychological disorders) | a |
| Ursodeoxycholic acid prescription for primary biliary cirrhosis, alcoholic cirrhosis, primary sclerosing cholangitis | **ATC code:** A05AA02 (ursodeoxycholic acid) **ICD-10 code:** K743 (primary biliary cirrhosis);  K703 K704 (alcoholic cirrhosis of liver); K830 (cholangitis); K80 (cholelithiasis) | a |
| Vitamin B12 medications for diabetic neuropathy | **ATC code:** B03BA04, B03BA05 (vitamin B12 drugs) **ICD-10 code:** E10, E11, E12, E13, E14 (diabetes mellitus); D51, D52, E538, E539 (vitamin B12 deficiency); G590, G632, E104, E107, E114, E117, E134, E137, E144 (diabetic neuropathy) | a |
| **Diagnostic or imaging tests (N=14)** |  |  |
| 1, 25-dihydroxyvitamin D testing in the absence of hypercalcemia or decreased kidney function | **Japanese Medical Practice Code:** 160158150 (vitamin D test)  **ICD-10 code:** N181, N182, N183 (CKD); E8352 (hypercalcemia); E211 (secondary hyperparathyroidism); D86 (sarcoidosis); J65, A15-A19, B90 (tuberculosis); C43, C44, C50, C56, C64, C65, C67, C81-C86, C88, C90-C96 (selected neoplasms) | a |
| Bone mineral density testing at frequent intervals | **Japanese Medical Practice Code:** 160091310, 160186870, 160147310, 160170410 (mineral density test)  **ICD-10 code:** M80, M81, M82 (osteoporosis) | a |
| Early imaging for acute low back pain | **Japanese Medical Practice Code:** 170001910, 170027910, 170028310, 170002410 (Plain x-ray); 170011810, 170028610, 170033410, 170034910, 170038710, 170038810, 170038910, 170039010, 170039110, 170040210, 170040310, 170040410, 170040610, 170040710, 170040810, 170041010, 170041110 (CT); 170015210, 170020110, 170033510, 170035010, 170041410, 170041510, 170041610, 170041710 (MRI).  **ICD-10 code**: M4306, M4309, M4316, M4317, M4319, M4329, M4646, M4649, M4716, M4719, M4786, M4789, M4799, M4800, M4806, M4808, M4809, M510, M511, M512, M513, M518, M519, M5326, M5328, M5329, M533, M539, M5416–M5419, M543, M544, M545, M548, M549, M961, M99, Q762, S33 (low back pain); S00-T98, B20-B24, D80-89, I33, A40, A41, J65, A15-A19, B90, M86, M511, M5416-M5419, R50, R53, D63, D64 (diagnoses warranting imaging [injuries, HIV disease, immune disorders, endocarditis, tuberculosis, osteomyelitis, intervertebral disk disorders and sciatica, fever, malaise/fatigue, and anemia]) | a  (Including costs for reading) |
| Electroencephalography (EEG) for headache | **Japanese Medical Practice Code:** 160075310, 160075750, 160075850, 160075950, 160076050, 160200510, 160170610, 160207510, 160187010 (EEG)  **ICD-10 code:** G43, G44, R51 (headache); A080, A081, A084, F445, G253, G40, G41, G433, G513, G934, R56 (epilepsy or convulsions) | a |
| Frequent pulmonary function test (PFT) for stable chronic obstructive pulmonary disease (COPD) | **Japanese Medical Practice Code:** 160062610, 160062710, 160062810, 160063010 (PFT) **ICD-10 code:** J41, J42, J45, J47, J6x, J84, C33, C34 (chronic respiratory diseases except COPD); J43, J44 (COPD)  **Billing Category code ^d^:** Low- or intermediate-risk surgical procedures include surgeries of the breast (K472-K476), colectomy (K719), cholecystectomy (K672), transurethral resection of the prostate (K841), hysterectomy (K872-K879), orthopedic surgeries including arthroscopy (besides hip and knee replacement) (K023-K144), corneal transplant (K259), cataract removal (K282), retinal detachment (K275, K276, K277, K279, K280, K281, K284), hernia repair (K633, K634), lithotripsy (K768), coronary artery bypass grafting (CABG) (K552), aneurysm repair (K560), thromboendarterectomy (K551, K609), percutaneous transluminal coronary angioplasty (PTCA) (K546-K549), and pacemaker insertion (K597-K599). (Low- or intermediate-risk non-cardiothoracic surgical procedures [shown above] plus CABG, aneurysm repair, thromboendarterectomy, PTCA, and pacemaker insertion) | a |
| Hypercoagulability testing for patients with deep vein thrombosis | **Japanese Medical Practice Code:** 160154350, 160164050 (anticardiolipin antibodies), 160169150, 160197610 (lupus anticoagulant), 160192310, 160114110, 160192210, 160124850 (Protein C and S activity and antigen), 160016010 (factor V test)  **ICD-10 code:** I802 (deep vein thrombosis); I260, I269 (pulmonary embolism) | a |
| Imaging for uncomplicated headache | **Japanese Medical Practice Code:** 170011810, 170028610, 170033410, 170034910, 170038710, 170038810, 170038910, 170039010, 170039110, 170040210, 170040310, 170040410, 170040610, 170040710, 170040810, 170041010, 170041110 (CT); 170015210, 170020110, 170033510, 170035010, 170041410, 170041510, 170041610, 170041710 (MRI).  **ICD-10 code:** G43, G44, R51 (headache);  A080, A081, A084, F445, G253, G40, G41, G433, G513, G934, R56 (diagnoses warranting imaging [epilepsy or convulsions]); I60-I69, G45, Z8673, Z8674, (diagnoses warranting imaging [cardiovascular disease]); S00-S09 (diagnoses warranting imaging [injuries]), R20, R26, R41, R47 (diagnoses warranting imaging [neurologic symptoms]), C00-D48 (diagnoses warranting imaging [cancer]) | a  (Including costs for reading) |
| Preoperative breast MRI | **Japanese Medical Practice Code:** 170035170 (breast MRI); 150121610, 150303110, 150316510, 150262710, 150121710, 150121810, 150121910, 150386510, 150345870, 150345970 (breast cancer surgery); 150121410, 150405810, 150413710 (surgery for benign breast diseases)  **ICD-10 code**: C50 (breast cancer) (C5001 for breast Paget’s disease); R798 (hereditary breast and ovarian cancer syndrome) | a  (Including healthcare costs for MRI imaging and reading and supplies, such as contrast media.) |
| Preoperative echocardiogram | **Japanese Medical Practice Code:** 160072510, 160072610 (echocardiogram); 111000570, 112001110, 112006470, 113016270, 113018570, 111000670, 112001210, 112006570, 113016370, 113018670, 111000770, 112001310, 112006670, 113016470, 113018770 (off-hour visit)  **Billing Category code ^d^:** Low- or intermediate-risk non-cardiothoracic surgical procedures include surgeries of the breast (K472-K476), colectomy (K719), cholecystectomy (K672), transurethral resection of the prostate (K841), hysterectomy (K872-K879), orthopedic surgeries including arthroscopy (besides hip and knee replacement) (K023-K144), corneal transplant (K259), cataract removal (K282), retinal detachment (K275, K276, K277, K279, K280, K281, K284), hernia repair (K633, K634), and lithotripsy (K768). | a |
| Preoperative PFT | **Japanese Medical Practice Code:** 160062610, 160062710, 160062810, 160063010 (PFT); 111000570, 112001110, 112006470, 113016270, 113018570, 111000670, 112001210, 112006570, 113016370, 113018670, 111000770, 112001310, 112006670, 113016470, 113018770 (off-hour visit).  **Billing Category code ^d^:** Low- or intermediate-risk surgical procedures include surgeries of the breast (K472-K476), colectomy (K719), cholecystectomy (K672), transurethral resection of the prostate (K841), hysterectomy (K872-K879), orthopedic surgeries including arthroscopy (besides hip and knee replacement) (K023-K144), corneal transplant (K259), cataract removal (K282), retinal detachment (K275, K276, K277, K279, K280, K281, K284), hernia repair (K633, K634), lithotripsy (K768), coronary artery bypass grafting (CABG) (K552), aneurysm repair (K560), thromboendarterectomy (K551, K609), percutaneous transluminal coronary angioplasty (PTCA) (K546-K549), and pacemaker insertion (K597-K599) | a |
| Preoperative stress testing or stress testing for stable coronary disease | **Japanese Medical Practice Code**: 160069210, 160069310, 160069410, 160069910, 160070050, 160198810, 170020070, 170027770, 170027870 (stress test); 111000570, 112001110, 112006470, 113016270, 113018570, 111000670, 112001210, 112006570, 113016370, 113018670, 111000770, 112001310, 112006670, 113016470, 113018770 (off-hour visit)  **ICD-10 code:** I25 (chronic ischemic heart disease)  **Billing Category code ^d^:** Low- or intermediate-risk non-cardiothoracic surgical procedures include surgeries of the breast (K472-K476), colectomy (K719), cholecystectomy (K672), transurethral resection of the prostate (K841), hysterectomy (K872-K879), orthopedic surgeries including arthroscopy (besides hip and knee replacement) (K023-K144), corneal transplant (K259), cataract removal (K282), retinal detachment (K275, K276, K277, K279, K280, K281, K284), hernia repair (K633, K634), and lithotripsy (K768) | c |
| PTH testing for patients with stage 1-3 chronic kidney disease | **Japanese Medical Practice Code:** 160035510 (PTH test); 140036710, 140051010, 140051110, 140057810, 140057910, 140058010, 140059310, 140059410, 140060210, 140060310, 140060410, 140059070, 140059170, 140052810, 140058110, 140058210, 140058410, 140058510, 140058610, 140007710, 140008170, 140052570, 140052970, 140007910, 140058770, 140058870, 140033770, 140058970, 140055970, 140029850, 140053670 (dialysis).  **ICD-10 code:** N181, N182, N183 (chronic kidney disease); E8352 (hypercalcemia) | a |
| Routine preoperative medical testing | **Japanese Medical Practice Code:** 150253010, 150280650, 150315610, 150356210, 150356310, 150380950, 150381050 (cataract surgery); 170001910, 170002410, 170021750, 170021850, 170023950, 170024050, 170027910, 170028310, 170031350, 170031450, 170031550, 170031650, 170032050, 170032150, 170032450, 170032550 (x-ray [photograph]); 170000410, 170000910, 170001250, 170001650, 170021550, 170021650, 170022730, 170023750, 170023850; 170000410, 170000910, 170001250, 170001650, 170021550, 170021650, 170022730, 170023750, 170023850 (x-ray [diagnosis]); 160068410, 160068510, 160068610 (electrocardiogram); 170001910, 170002410, 170021750, 170021850, 170023950, 170024050, 170027910, 170028310, 170031350, 170031450, 170031550, 170031650, 170032050, 170032150, 170032450, 170032550 (x-ray [photograph]); 170000410, 170000910, 170001250, 170001650, 170021550, 170021650, 170022730, 170023750, 170023850; 170000410, 170000910, 170001250, 170001650, 170021550, 170021650, 170022730, 170023750, 170023850 (x-ray (diagnosis)); 160068410, 160068510, 160068610 (electrocardiogram)  **Billing Category code ^d^:** D005-D016 (blood test). Low- or intermediate-risk non-cardiothoracic surgical procedures include surgeries of the breast (K472-K476), colectomy (K719), cholecystectomy (K672), transurethral resection of the prostate (K841), hysterectomy (K872-K879), orthopedic surgeries including arthroscopy (besides hip and knee replacement) (K023-K144), corneal transplant (K259), cataract removal (K282), retinal detachment (K275, K276, K277, K279, K280, K281, K284), hernia repair (K633, K634), and lithotripsy (K768) | a |
| Serum T3 level testing for patients with hypothyroidism | **Japanese Medical Practice Code:** 160031310, 160033210 (T3 test)  **ICD-10 code:** E02, E03 (hypothyroidism) | a |
| **Procedures (N=18)** |  |  |
| Arthroscopic surgery for knee osteoarthritis | **Japanese Medical Practice Code:** 150309510, 150310410, 150311310, 150312410  **ICD-10 code:** M150, M153, M159, M17, M1909, M1919, M1929, M1999 (osteoarthritis); M224, M942 (chondromalacia); M232, S832 (meniscal tear) | b |
| Artificial liver support for acute liver failure | **Japanese Medical Practice Code:** 140008410 (Artificial liver support)  **ICD-10 code:** B150, B162, B171, B172, B190, K711, K720, K729 (acute liver failure); B181, B182, B189, K700, K701, K702, K703, K704, K709, K713, K717, K721, K730, K732, K738, K739, K740, K741, K743, K744, K745, K746, K754, K758, K760, K761 (chronic or subacute liver disease) | a |
| Bed rest for threatened preterm labor | **ATC code:** H02AB01, H02AB02, H02AB03, H02AB04, H02AB05, H02AB06, H02AB07, H02AB08, H02AB09, H02AB10, H02AB11, H02AB12, H02AB13, H02AB14, H02AB15 (systemic corticosteroids); G02CX, C08CA (tocolytic drugs) **ICD-10 code:** O (pregnancy-related conditions); O600 (threatened preterm labor); O42 (premature rupture of membranes); O15 (eclampsia); O142 (HELLP syndrome); O30 (multiple pregnancy); O13 (hypertensive disorders of pregnancy); O249, E10, E11, E12, E13, E14 (diabetes); O46 (antepartum hemorrhage); O41 (other disorders of amniotic fluid and membranes); O45 (placental abruption); O44 (placenta previa) | b |
| Carotid endarterectomy in asymptomatic patients | **Japanese Medical Practice Code:** 150322710 (carotid endarterectomy); 140036710, 140051010, 140051110, 140057810, 140057910, 140058010, 140059310, 140059410, 140060210, 140060310, 140060410, 140059070, 140059170, 140052810, 140058110, 140058210, 140058410, 140058510, 140058610, 140007710, 140008170, 140052570, 140052970, 140007910, 140058770, 140058870, 140033770, 140058970, 140055970, 140029850, 140053670 (dialysis)  **ICD-10 code:** H340-H342 (retinal artery occlusion); G45 (TIA); I60-I63, I66 (stroke); R25, R430-R432, R270, R278, R279, R290, R291, R295, R683, R414, R471, R478, R20 (nervous and musculoskeletal symptoms); I21-I23 (acute myocardial infarction); J43, J44 (chronic obstructive pulmonary disease); alcohol related disorder (F10); presence of cardiac pacemaker (Z950) | b |
| Emergency surgery for acute appendicitis shortly after admission (<24 h) | **Japanese Medical Practice Code:** 150181610,150337610 (appendectomy); 150000490, 150371390, 150000690, 150371490 (additional fee for after-hours emergency surgery) **ICD-10 code:** K358 (acute appendicitis without peritonitis); K352, K353 (appendicitis with peritonitis); K65 (peritonitis) | a  (Including only the healthcare cost associated with the Japanese Medical Practice Code for additional fee for emergency surgery.) |
| Endoscopy for dyspepsia for people < 55 years or colonoscopy for constipation in people < 50 years | **Japanese Medical Practice Code:** 160093810 (esophagogastroduodenoscopy), 160094710, 160094810, 160094910, 160202750 (colonoscopy)  **ICD-10 code:** F453, K30, R101 (dyspepsia) K589, K590 (constipation); D50-53, D55-64 (anemia), R131 (dysphagia), R634, R64 (weight loss); C15-26, C784-788 (cancer of digestive system), K20-K31, K35-K38, K40-K44, K50-K52, K55-K64 (except for K589 and K590), K65-K67, K70-K77, K80-K87, K90-K93 (other diseases of the digestive system) | c |
| Endotoxin apheresis for sepsis | **Japanese Medical Practice Code:** 140037250, 140061610 (endotoxin apheresis) | a |
| Feeding tube for severe dementia | **Japanese Medical Practice Code:** 150171610, 150380510 (gastrostomy); 150362110 (percutaneous trans-esophageal gastro-tube insertion), 140054710 (elemental diet tube insertion); 114004310, 114045510, 140023210, 140023350 (tube feeding management); 114000110, 114001110, 114007610, 114007710, 114012910, 114018010, 114019510, 114019610, 114019710, 114019810, 114030310 (home care); 114035510, 114035610, 114035710, 114035810, 114035910, 114036010, 114036110, 114036210, 114036310, 114036410, 114036510, 114036610, 114036710, 114036810, 114036910, 114037010, 114037110, 114037210, 114037310, 114037410, 114037510, 114037610, 114037710, 114037810, 114037910, 114038010, 114038110, 114038210, 114038310, 114038410, 114038510, 114038610, 114038710, 114038810, 114038910, 114039010, 114041870, 114042110, 114042210, 114042810, 114046310, 114058010, 114058110, 114058210, 114058310, 114058410, 114058510, 114058610, 114058710, 114058910, 114059110, 114059210, 114059310, 114059410, 114059510, 114059610, 114059910 (institutional care).  **ICD-10 code:** F00-F03 G30, B220 (dementia); K50 (Crone’s disease); K51 (ulcerative colitis); K221 (ulcer of the esophagus); K222 (esophageal obstruction); K223 (perforation of esophagus); T902, T20, S07 (head injury and burn); Q39 (congenital malformations of esophagus); G (excluding G30) (diseases of nervous system other than dementia) | a  (The cost of tube feeding meals was not included) |
| Inferior vena cava (IVC) filters for the prevention of pulmonary embolism | **Japanese Medical Practice Code:** 150263510 (IVC filter insertion)  **ICD-10 code:** I260, I269 (pulmonary embolism); I802 (deep vein thrombosis) | a |
| Percutaneous coronary intervention (PCI) with angioplasty or stent placement for stable coronary disease | **Japanese Medical Practice Code:** 150375110, 150375410 (PCI, not related to acute coronary syndrome); 111000570, 112001110, 112006470, 113016270, 113018570, 111000670, 112001210, 112006570, 113016370, 113018670, 111000770, 112001310, 112006670, 113016470, 113018770 (off-hour visit)  **ICD-10 code:** I20-I25 (ischemic heart disease or angina) (I21-I23 for acute myocardial infarction) | b  (In the case of outpatient PCI, all costs incurred on the same day of service were included in healthcare spending estimates. ) |
| Pulmonary artery catheterization (PAC) in the intensive care unit (ICU) | **Japanese Medical Practice Code:** 160183910, 160075010, 160075170 (pulmonary artery catheterization);  **Billing Category code ^d^:** A300, A301 (ICU): K00x-K91x (surgical procedures).  **ICD-10 code:** I27 (pulmonary hypertension); I319 (cardiac tamponade) | a |
| Renal angioplasty | **Japanese Medical Practice Code:** 150152010  **ICD-10 code:** I150 (renovascular hypertension), I701 (renal atherosclerosis); I773 (fibromuscular dysplasia) | b |
| Spinal fusion for lumber stenosis | **Japanese Medical Practice Code:** 150314810, 150282510, 150282610, 150314610, 150314710, 150368870, 150368970, 150369070, 150314810, 150369170, 150369370, 150369170, 150397210, 150314210 (spinal fusion)  **ICD-10 code:** M4806 (lumber stenosis); M431 (spondylolisthesis); M796 (pain in foot); M511, M5416-M5419 (radiculopathy); M543, M544 (sciatica); Q76 (congenital malformations of spine) | a  (The additional healthcare spending [including the healthcare spending on materials] compared to the healthcare spending of laminectomy was used as an estimate of the healthcare spending.) |
| Spinal injection for low back pain | **Japanese Medical Practice Code:** 150235510, 150236010, 150242110, 150266010, 150265010, 150235710, 150350710, 150236010, 150265710, 150351310, 150239110  **ICD-10 code:** M4306, M4309, M4316, M4317, M4319, M4329, M4646, M4649, M4716, M4719, M4786, M4789, M4799, M4800, M4806, M4808, M4809, M510, M511, M512, M513, M518, M519, M5326, M5328, M5329, M533, M539, M5416–M5419, M543, M544, M545, M548, M549, M961, M99, Q762, S33 (back pain); M511, M5416-M5419 (radiculopathy) | c |
| Surgery for vesicoureteral reflux | **Japanese Medical Practice Code:** 150201950, 150326310, 150365410  **ICD-10 code:** N137, Q627 (vesicoureteral reflux) | b |
| Traction therapy for back pain or neck pain | **Japanese Medical Practice Code:** 140048010 (traction therapy)  **ICD-10 code:** M4306, M4309, M4316, M4317, M4319, M4329, M4646, M4649, M4716, M4719, M4786, M4789, M4799, M4800, M4806, M4808, M4809, M510, M511, M512, M513, M518, M519, M5326, M5328, M5329, M533, M539, M5416–M5419, M543, M544, M545, M548, M549, M961, M99, Q762, S33 (back pain); M50, M540, M5411, M542 (neck pain); C412 (malignant spinal tumor); M05, M06, M790 (rheumatoid arthritis); M45, M46 (spondylitis); M80, M81, M82 (osteoporosis) | a |
| Vertebroplasty for osteoporotic vertebral fractures | **Japanese Medical Practice Code:** 150355210 (vertebroplasty)  **ICD-10 code:** M8008, M8018, M8028, M8038, M8048, M8058, M8088, M8098, M48.4, M84.4, S12, S220, S221, S320, T08, Y427 (vertebral fracture); C412, C795 (bone cancers); C90 (myeloma); D180 (hemangioma) | b |

^a^ Including only the healthcare spendings associated with the Japanese Medical Practice Codes or ATC codes.

^b^ All costs incurred in the same hospitalization were included in healthcare spending estimates. Note that vertebroplasty, renal artery angioplasty, and arthroscopic knee surgery are exclusively conducted in inpatient settings in Japan, unlike in the United States.

^c^ All costs incurred on the same day of service were included in healthcare spending estimates.

^d^ The Billing Category code represents a broad category that groups together multiple medical procedures of the same type. For example, Billing Category code K511 (pneumonectomy) includes wedge resection (Japanese Medical Practice Codes 150129710), segmental resection (less than one lung lobe) (150129810), lobectomy (150129910), composite resection (more than one lung lobe) (150130010), total pneumonectomy on one side (150130110), and pneumonectomy with brachioplasty (150317110).

**eTable 3. Low-value care items excluded due to being unmeasurable using claims data**

| **Unmeasurable low-value items** | **Reason for immeasurability** |
| --- | --- |
| Stool culture for inpatients with diarrhea, performed ≥3 days after admission | The coding system only covers samples from the digestive tract, lacking specific codes for stool culture. |
| Sterile glove use for laceration repair performed in the emergency department | No codes are available for documenting the use of sterile gloves. |
| Contrast-enhanced chest CT in patients with a low pretest probability of pulmonary embolism (PE) | Clinical information to identify low probability of PE (such as Well’s score) is lacking in claims data. |
| Routine chest radiography in patients without respiratory symptoms | Clinical information to identify respiratory symptoms is lacking in claims data. |
| Repeated and long-term chest CT scans for patients with lung nodules | Information on pulmonary nodules cannot be identified from claims data. |
| Chest CT screening in patients at low risk for lung cancer | Lack of smoking cessation information; furthermore, preventive care is not included in claims data. |
| Use of pulmonary vasodilators approved only for Group 1 pulmonary hypertension in patients with Group 2 or 3 pulmonary hypertension | Insufficient accuracy in group classification of pulmonary hypertension using claims data. |
| Continuation of home oxygen therapy without reassessment in patients who initiated home oxygen therapy (HOT) for an acute illness | Since SpO2 values cannot be obtained from claims data and there is no information on whether the attending physician assessed the need for HOT, identification using claims data is challenging. |
| Screening for ovarian cancer in asymptomatic women using CA-125 testing and transvaginal ultrasonography | Claims data do not include preventive care. |
| Cervical cytology sampling with a cotton swab in non-pregnant women | There are no existing codes for the use of cotton swabs. |
| Bed rest after lumbar puncture | There are no existing codes for bed rest after lumbar puncture. |
| Topical antibiotics for prevention of surgical site infection after dermatologic surgery | Wound status data are not available in claims data. |
| No medication adjustment (deprescribing) performed in frail elderly individuals | Frailty status cannot be identified, and the cost associated with failure to deprescribe cannot be measured. |
| Erythropoietin therapy aimed at maintaining hemoglobin >10 g/dL | Blood test information at the visit is not available in claims databases. |
| Activated charcoal administration for acute alcohol intoxication | Use of activated charcoal cannot be identified from claims data. |
| Routine intravenous corticosteroid use in patients hospitalized for asthma or chronic obstructive pulmonary disease exacerbations | Given the equivalent efficacy and similar costs of oral versus intravenous steroids in mild-to-moderate asthma or COPD exacerbation admissions, these cases are excluded from evaluation. |
| Routine oxygen administration for acute illnesses targeting high oxygen saturation levels | Peripheral oxygen saturation values are unavailable. |
| Levothyroxine for subclinical hypothyroidism in older adults | Identification in claims data is challenging due to the absence of diagnosis codes for subclinical hypothyroidism and lack of laboratory data. |
| PSA screening at age >75 years | Preventive care services are generally not covered in public insurance; therefore, PSA screening cannot be reliably identified from claims data. Although a substantial number of PSA tests are reportedly conducted under insurance coverage, claims data do not differentiate between PSA tests performed for diagnostic purposes and those performed as routine screening. |
| Bacteriuria screening for asymptomatic patients | Clinical information to identify symptoms such as frequent urination and discomfort during urination is unavailable in claims data. |

**eTable 4. Volume and spending of 52 low-value care services, using narrower definitions**

|  | Price (thousand JPY) ^a^ | Sample low-value care estimates | | National low-value care estimates ^b^ | |
| --- | --- | --- | --- | --- | --- |
|  |  | Volume | Spending (million JPY) | Volume (thousand) | Spending (billion JPY) |
| **Price category, very low** ^a^ | | | | | |
| Topical salicylates or long-term topical nonsteroidal anti-inflammatory drugs (NSAIDs) therapy for chronic pain | 0.9 | 1,738,631 | 1532.3 | 52,611.7 | 45.6 |
| Antibiotics for acute respiratory infections (ARIs) | 0.7 | 182356 | 121.5 | 15,282.8 | 9.9 |
| Antihistamines for ARIs | 0.5 | 85250 | 45.8 | 7,961.3 | 3.2 |
| Expectorants for ARIs | 0.2 | 271645 | 43.9 | 25,903.6 | 3.6 |
| Preoperative pulmonary function test (PFT) | 1.0 | 12664 | 12.1 | 573.0 | 0.5 |
| Antihistamine-analgesic combinations for ARIs | 0.1 | 88347 | 10.7 | 4,963.6 | 0.6 |
| Traction therapy for back or neck pain | 0.4 | 28861 | 10.1 | 1,060.7 | 0.4 |
| Codeine for ARIs | 0.2 | 39643 | 8.7 | 2,871.7 | 0.6 |
| Oral betamimetics | 0.8 | 4360 | 3.6 | 594.4 | 0.5 |
| Antibiotics prescription for uncomplicated colon diverticulitis | 0.6 | 1412 | 0.9 | 75.2 | 0.05 |
| Dextromethorphan prescription for ARIs in children (<18 years) | 0.1 | 9041 | 0.9 | 996.9 | 0.1 |
| Tranexamic acid administration for intracerebral hemorrhage | 0.3 | 910 | 0.3 | 33.2 | 0.01 |
| Vitamin B12 medications for diabetic neuropathy | 0.7 | 309 | 0.2 | 12.3 | 0.01 |
| Tricyclic antidepressants prescription for children | 0.4 | 123 | 0.05 | 12.4 | 0.004 |
| Antibiotics prescription for threatened preterm labor | 0.6 | 42 | 0.02 | 5.7 | 0.003 |
| **Price category, low** ^a^ | | | | | |
| Back imaging | 3.3 | 211914 | 693.7 | 9819.4 | 31.6 |
| Injection for low back pain | 6.0 | 74125 | 444.0 | 2,042.4 | 12.7 |
| Short-term repeat bone mineral density testing | 1.5 | 148452 | 221.9 | 4,809.3 | 7.2 |
| Pregabalin for low back pain | 1.7 | 84701 | 140.6 | 2,761.8 | 4.6 |
| Preoperative medical testing | 5.4 | 18432 | 99.2 | 613.5 | 3.5 |
| Serum T3 testing for hypothyroidism | 1.2 | 72301 | 89.5 | 2,892.6 | 3.6 |
| Preoperative echocardiogram | 8.8 | 7118 | 62.6 | 247.5 | 2.2 |
| Neuraminidase for non-severe influenza | 2.9 | 7879 | 23.0 | 785.9 | 2.4 |
| 1, 25-dihydroxy vitamin D  testing in the absence of  hypercalcemia or decreased  kidney function | 3.9 | 1588 | 6.2 | 62.7 | 0.2 |
| Electroencephalography for headache | 7.1 | 862 | 6.1 | 57.4 | 0.4 |
| Hypercoagulability testing for patients with deep vein thrombosis (DVT) | 2.2 | 2288 | 5.1 | 104.1 | 0.2 |
| Antibiotics prescription for acute pancreatitis | 3.7 | 968 | 3.5 | 31.9 | 0.1 |
| Non-preoperative frequent PFT for stable chronic obstructive pulmonary disease (COPD) | 2.0 | 615 | 1.2 | 20.8 | 0.04 |
| Ursodeoxycholic acid prescription for primary biliary cirrhosis, alcoholic cirrhosis, primary sclerosing cholangitis | 1.2 | 548 | 0.7 | 23.1 | 0.03 |
| Pulmonary artery catheter in the intensive care unit | 6.3 | 52 | 0.3 | 2.7 | 0.02 |
| Nitroglycerin or isosorbide dinitrate for perioperative management of non-cardiac surgery | 1.6 | 116 | 0.2 | 4.5 | 0.01 |
| Parathyroid hormone (PTH) test for patients with stages 1–3 chronic kidney disease | 1.7 | 9 | 0.01 | 0.4 | 0.001 |
| Intravenous anti-herpes drugs for sudden sensorineural hearing loss | 7.1 | 1 | 0.01 | 0.1 | 0.001 |
| **Price category, medium** ^a^ | | | | | |
| Head imaging | 15.4 | 19477 | 299.9 | 1102.0 | 17.1 |
| Preoperative stress testing or stress testing for stable coronary disease | 31.6 | 4253 | 134.6 | 161.8 | 4.9 |
| Endoscopy for dyspepsia for  people < 55 years or  colonoscopy for constipation  in people < 50 years | 22.7 | 1685 | 38.3 | 163.4 | 3.7 |
| Feeding tube for severe dementia | 76.9 | 182 | 14.0 | 4.2 | 0.3 |
| Preoperative breast magnetic resonance imaging (MRI) | 27.5 | 216 | 5.9 | 10.6 | 0.3 |
| Intravenous betamimetics, >48h | 11.5 | 370 | 4.3 | 50.6 | 0.6 |
| IV sivelestat for acute respiratory distress syndrome (ARDS) | 27.0 | 147 | 4.0 | 4.3 | 0.1 |
| Artificial liver support for acute liver failure | 20.0 | 42 | 0.8 | 1.7 | 0.03 |
| Emergency surgery for acute appendicitis shortly after admission (<24 h) | 62.7 | 7 | 0.4 | 0.5 | 0.03 |
| **Price category, high** ^a^ | | | | | |
| Vertebroplasty for osteoporotic vertebral fractures | 1463.8 | 410 | 600.2 | 10.3 | 15.3 |
| Percutaneous coronary intervention (PCI) for stable coronary disease | 1022.6 | 242 | 247.5 | 9.4 | 10.3 |
| Spinal fusion for lumber stenosis | 243.6 | 644 | 156.9 | 25.9 | 6.5 |
| Arthroscopic surgery for knee osteoarthritis | 705.3 | 98 | 69.1 | 5.8 | 3.7 |
| Bed rest for threatened preterm labor | 517.7 | 94 | 48.7 | 13.4 | 6.6 |
| Carotid endarterectomy in asymptomatic patients | 1262.7 | 37 | 46.7 | 1.1 | 1.4 |
| Endotoxin apheresis for sepsis | 643.9 | 47 | 30.3 | 1.6 | 1.0 |
| Renal angioplasty | 1756.9 | 17 | 29.9 | 0.6 | 1.1 |
| Inferior vena cava (IVC) filters for prevention of pulmonary embolism | 101.6 | 86 | 8.7 | 2.6 | 0.3 |
| Surgery for vesicoureteral reflux | 815.8 | 1 | 0.8 | 0.1 | 0.1 |
| **Total** | - | 3123618 | 5329.8 | 138808.6 | 207.2 |

Low-value care measures were grouped by price categories, and within each category, the measures were ordered by spending in the analytic sample.

^a^ Price was defined as average spending per service.

^b^ National extrapolation was adjusted for age, sex, and regions.

**eTable 5. Volume and cost of low-value care services in fiscal year 2022 overall and by age group, using broader low-value care definitions**

|  | **No. of beneficiaries** | **Volume** | | | **Spending** | | |
| --- | --- | --- | --- | --- | --- | --- | --- |
|  |  | Total LVC volume, no. of services | LVC volume per 1,000 beneficiaries | No. (%) of beneficiaries receiving LVC | Total LVC cost, million JPY (%) | LVC cost per 1,000 beneficiaries (million JPY) | % of total healthcare spending within each category ^a^ |
| **Overall** | 1,923,484 | 3,714,442 | 1931.1 | 760,101 (39.5) | 8403.1 | 4.37 | 1.02 |
| **Stratified by age category** |  |  |  |  |  |  |  |
| <18 years | 162,563 | 307,740 | 1893.1 | 84,053 (51.7) | 123.2 | 0.76 | 0.57 |
| 18–64 years | 784,945 | 684,492 | 872.0 | 223,135 (28.4) | 1445.7 | 1.84 | 0.80 |
| 65–74 years | 402,795 | 728,403 | 1808.4 | 152,271 (37.8) | 2006.6 | 4.98 | 0.96 |
| ≥75 years | 573,181 | 1,993,807 | 3478.5 | 300,642 (52.5) | 4827.5 | 8.42 | 1.17 |

^a^ Total healthcare spendings (denominators) were 823.7 billion, 21.5 billion, 180.9 billion, 209.8 billion, and 411.6 billion JPY for the analytic sample overall, aged <18 years, aged 18–64 years, 65–74 years, and ≥75 years, respectively.

**eTable 6. Volume and spending of 52 low-value care service, using broader low-value care definitions**

|  | Price (thousand JPY) ^a^ | Sample low-value care estimates | | National low-value care estimates ^b^ | |
| --- | --- | --- | --- | --- | --- |
|  |  | Volume | Spending (million JPY) | Volume (thousand) | Spending (billion JPY) |
| **Price category, very low** ^a^ | | | | | |
| Topical salicylates or long-term topical nonsteroidal anti-inflammatory drugs (NSAIDs) therapy for chronic pain | 0.9 | 1738631 | 1532.3 | 52611.7 | 45.6 |
| Antibiotics for acute respiratory infections (ARIs) | 0.7 | 207846 | 143.3 | 17570.2 | 11.9 |
| Antihistamines for ARIs | 0.5 | 197376 | 101.7 | 19021.0 | 8.4 |
| Expectorants for ARIs | 0.2 | 312564 | 50.8 | 29972.0 | 4.2 |
| Traction therapy for back or neck pain | 0.4 | 39523 | 13.8 | 1343.9 | 0.5 |
| Preoperative pulmonary function test (PFT) | 1.0 | 12684 | 12.1 | 573.8 | 0.5 |
| Antihistamine-analgesic combinations for ARIs | 0.1 | 89434 | 10.8 | 5038.5 | 0.6 |
| Codeine for ARIs | 0.2 | 45595 | 10.2 | 3367.3 | 0.7 |
| Oral betamimetics | 0.8 | 4360 | 3.6 | 594.4 | 0.5 |
| Vitamin B12 medications for diabetic neuropathy | 0.7 | 3650 | 2.4 | 121.3 | 0.1 |
| Dextromethorphan prescription for ARIs in children (<18 years) | 0.1 | 10457 | 1.0 | 1155.1 | 0.1 |
| Antibiotics prescription for uncomplicated colon diverticulitis | 0.6 | 1412 | 0.9 | 75.2 | 0.05 |
| Tricyclic antidepressants prescription for children | 0.5 | 523 | 0.3 | 55.2 | 0.03 |
| Tranexamic acid administration for intracerebral hemorrhage | 0.3 | 910 | 0.3 | 33.2 | 0.01 |
| Antibiotics prescription for threatened preterm labor | 0.6 | 61 | 0.04 | 8.6 | 0.01 |
| **Price category, low** ^a^ | | | | | |
| Back imaging for low back pain | 3.3 | 255415 | 830.5 | 11665.9 | 36.9 |
| Pregabalin for low back pain | 1.8 | 366749 | 660.9 | 12364.8 | 22.1 |
| Injection for low back pain | 6.0 | 75870 | 458.9 | 2098.7 | 13.2 |
| Preoperative medical testing | 7.2 | 57133 | 411.7 | 2424.0 | 17.6 |
| Short-term repeat bone mineral density testing | 1.5 | 159793 | 234.9 | 5275.4 | 7.7 |
| Serum T3 testing for hypothyroidism | 1.2 | 72301 | 89.5 | 2892.6 | 3.6 |
| Preoperative echocardiogram | 8.8 | 7149 | 62.9 | 248.7 | 2.2 |
| Neuraminidase for non-severe influenza | 2.9 | 7961 | 23.2 | 792.9 | 2.4 |
| Electroencephalography for headache | 7.1 | 519 | 3.7 | 63.6 | 0.5 |
| 1, 25-dihydroxy vitamin D testing in the absence of hypercalcemia or decreased kidney function | 3.9 | 1588 | 6.2 | 62.7 | 0.2 |
| Antibiotics prescription for acute pancreatitis | 3.9 | 1532 | 6.0 | 50.5 | 0.2 |
| Hypercoagulability testing for patients with deep vein thrombosis (DVT) | 2.2 | 2288 | 5.1 | 104.1 | 0.2 |
| Non-preoperative frequent PFT for stable chronic obstructive pulmonary disease (COPD) | 2.0 | 719 | 1.4 | 24.7 | 0.05 |
| Ursodeoxycholic acid prescription for primary biliary cirrhosis, alcoholic cirrhosis, primary sclerosing cholangitis | 1.2 | 571 | 0.7 | 23.7 | 0.03 |
| Pulmonary artery catheter in the intensive care unit | 6.3 | 52 | 0.3 | 2.7 | 0.02 |
| Nitroglycerin or isosorbide dinitrate for perioperative management of non-cardiac surgery | 1.9 | 168 | 0.3 | 5.9 | 0.01 |
| Intravenous anti-herpes drugs for sudden sensorineural hearing loss | 9.2 | 7 | 0.1 | 0.3 | 0.002 |
| Parathyroid hormone (PTH) test for patients with stages 1–3 chronic kidney disease | 1.7 | 9 | 0.01 | 0.4 | 0.001 |
| **Price category, medium**^a^ | | | | | |
| Head imaging | 15.5 | 25298 | 392.8 | 1327.6 | 20.7 |
| Endoscopy for dyspepsia for people < 55 years or colonoscopy for constipation in people < 50 years | 32.8 | 4368 | 143.4 | 417.6 | 13.5 |
| Preoperative stress testing or stress testing for stable coronary disease | 31.7 | 4254 | 134.7 | 161.8 | 4.9 |
| Feeding tube for severe dementia | 58.3 | 531 | 31.0 | 13.6 | 0.8 |
| Preoperative breast magnetic resonance imaging (MRI) | 27.6 | 240 | 6.6 | 11.8 | 0.3 |
| Intravenous betamimetics, >48h | 11.5 | 370 | 4.3 | 50.6 | 0.6 |
| IV sivelestat for acute respiratory distress syndrome (ARDS) | 27.0 | 147 | 4.0 | 4.3 | 0.1 |
| Artificial liver support for acute liver failure | 20.0 | 44 | 0.9 | 1.9 | 0.04 |
| Emergency surgery for acute appendicitis shortly after admission (<24 h) | 88.0 | 10 | 0.9 | 0.8 | 0.1 |
| **Price category, high** ^a^ | | | | | |
| Percutaneous coronary intervention (PCI) for stable coronary disease | 995.5 | 1847 | 1838.7 | 63.4 | 64.0 |
| Vertebroplasty for osteoporotic vertebral fractures | 1471.4 | 412 | 606.2 | 10.4 | 15.4 |
| Spinal fusion for lumber stenosis | 251.3 | 1042 | 261.9 | 42.4 | 10.6 |
| Arthroscopic surgery for knee osteoarthritis | 670.4 | 153 | 102.6 | 8.7 | 5.8 |
| Bed rest for threatened preterm labor | 626.2 | 115 | 72.0 | 16.3 | 9.2 |
| Carotid endarterectomy in asymptomatic patients | 1262.7 | 37 | 46.7 | 1.1 | 1.4 |
| Endotoxin apheresis for sepsis | 643.9 | 47 | 30.3 | 1.6 | 1.0 |
| Renal angioplasty | 1756.9 | 17 | 29.9 | 0.6 | 1.1 |
| Inferior vena cava (IVC) filters for prevention of pulmonary embolism | 101.6 | 86 | 8.7 | 2.6 | 0.3 |
| Surgery for vesicoureteral reflux | 386.3 | 11 | 4.2 | 1.3 | 0.4 |
| **Total** | - | 371442 | 8403.1 | 171781.43 | 330.5 |

Low-value care measures were grouped by price categories, and within each category, the measures were ordered by spending in the analytic sample.

^a^ Price was defined as average spending per service.

^b^ National extrapolation was adjusted for age, sex, and regions.

**eFigure 1. Proportion of total low-value care (LVC) volume and spending by price category, using broader low-value care definitions**

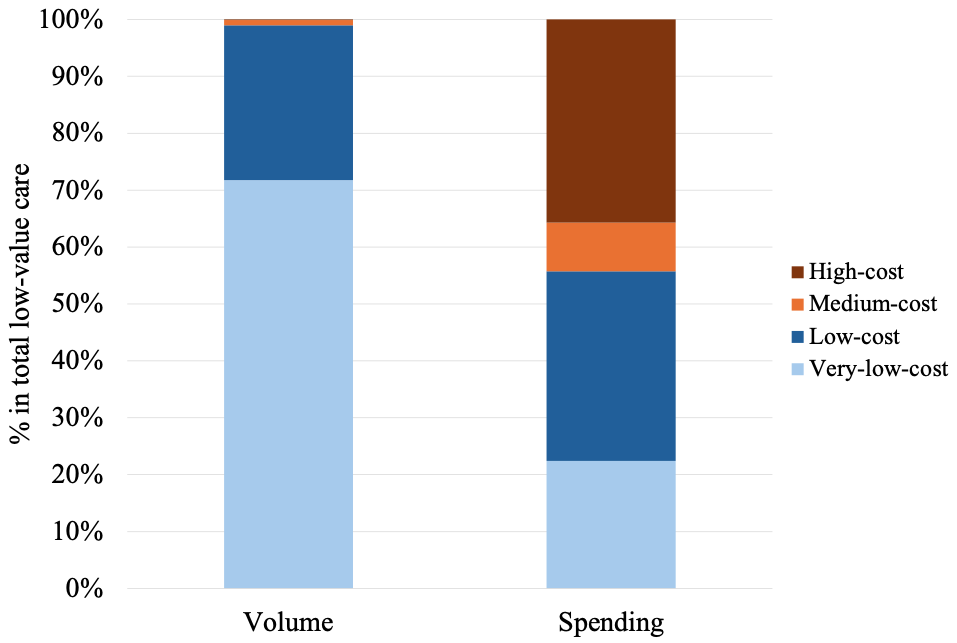

In this sensitivity analysis, we used the broader definition of LVC instead of the narrower one and examined the price category distribution of LVC services. Low-value care services were classified into four price categories according to an average spending per service: very-low-cost (under 1,000 Japanese yen [JPY; 1,000 JPY=8 US dollars in 2022]), low-cost (1,000 to 9,999 JPY), medium-cost (10,000 to 99,999 JPY), and high-cost (100,000 JPY or more). Categories of very-low-cost, low-cost, medium-cost, and high-cost consisted of 15, 18, 9, and 10 services, respectively.

The volume of very-low or low-cost services accounted for 98.9% of the volume of LVC services, and the total spending on these services (4.7 billion JPY [37.5 million USD], 55.7% of total unnecessary spending of 8.4 billion JPY [67.2 million USD]) exceeded the total spending on medium- or high-cost services (3.7 billion JPY [29.8 million USD], 44.3% of total unnecessary spending).

**eFigure 2. Proportion of total low-value care (LVC) volume and spending by price category, using alternative price category cutoffs**

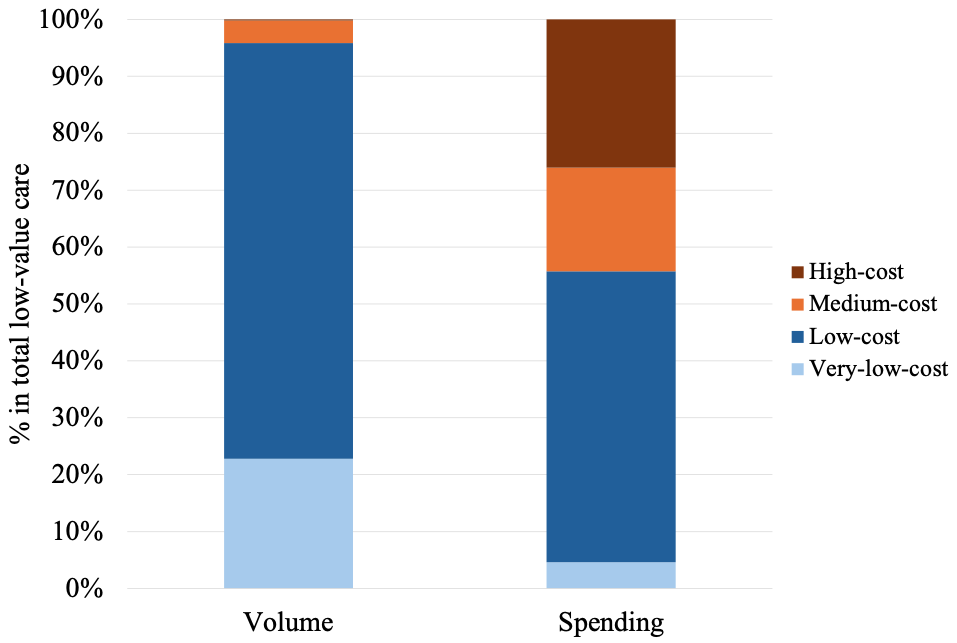

In this sensitivity analysis, we classified low-value care services into quartiles according to an average spending per service: very-low-cost (under 825 Japanese yen [JPY], equivalent to 5.5 US dollars [USD]), low-cost (825 JPY [5.5 USD] to 3,750 JPY [25 USD]), medium-cost (3,750 JPY [25 USD] to 30,000 JPY [200 USD]), and high-cost (30,000 JPY [200 USD] or more). Categories of very-low-cost, low-cost, medium-cost, and high-cost consisted of 13, 13, 13, and 13 services, respectively.

The volume of very-low-cost or low-cost services accounted for 95.8% of the volume of LVC services, and the total spending on these services (3.0 billion JPY [23.8 million USD], 55.7% of total unnecessary spending of 5.3 billion JPY [42.6 million USD]) far exceeded the total spending on medium- or high-cost services (2.4 billion JPY [18.9 million USD], 44.3% of total unnecessary spending).

**eFigure 3. Proportion of total low-value care (LVC) volume and spending by price category, based on national extrapolation that were age-, sex-, and region-adjusted.**

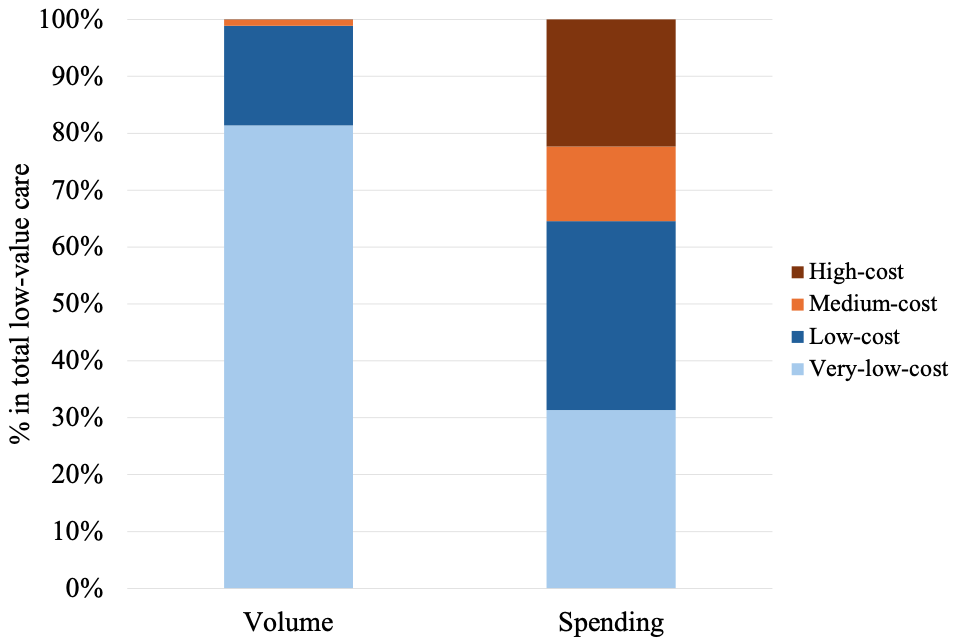

In this sensitivity analysis, we reanalyzed cost category distributions of LVC services based on national volume/spending estimates (instead of the convenient sample estimates) derived by extrapolating age-specific per capita volume/spending to the national population.

When extrapolated to the national population, the volume of very-low-cost or low-cost services accounted for 98.9% of the total volume of LVC services (137.2 million/138.8 million times), and the total spending on these services (133.8 billion JPY [1.1 billion USD], 64.6% of total unnecessary spending of 207.2 billion JPY [1.7 billion USD]) far exceeded the total spending on medium- or high-cost services (73.5 billion JPY [0.6 billion USD], 35.4% of total unnecessary spending).
